# Impact of PCSK9 inhibitors on bone disease: A comprehensive drug-target Mendelian randomization study

**DOI:** 10.1101/2025.03.09.25323603

**Authors:** Wangyu Yang, Xiaohui Wang, Hao Yang, Dingjun Hao

**Author notes:** Correspondence: **Hao Yang**, **Dingjun Hao**.

## Abstract

**Background:** While certain studies suggest a relationship between hyperlipidemia and bone metabolism, the exact nature of proprotein convertase subtilisin/kexin 9 inhibitors (PCSK9i, targeted by, eg, alirocumab), which were originally developed for lowering LDL cholesterol inhibitors, with bone disease is still unclear. This research endeavors to uncover the potential causal relationship between PCSK9i and several most popular bone diseases (osteoporosis (OP), osteoarthritis (OA), rheumatoid osteoarthritis (RA)) using Mendelian randomization (MR).

**Methods:** This study employed a comprehensive approach involving the extraction of single-nucleotide polymorphisms (SNPs) from genome-wide association studies (GWAS), followed by rigorous quality checks. PCSK9i instrumental variables were utilized to evaluate the effect of cholesterol-lowering drugs on osteoporosis, osteoarthritis, and rheumatoid arthritis.

**Results:** PCSK9i instrumental variables were validated using familial combined hyperlipidemia summary data. Our analysis did not reveal a significant causal relationship between PCSK9i and OP. However, there was an observed an increased Lumbar-spine bone mineral density (LS-BMD) with PCSK9i intaking (OR = 1.157, 95% CI: 0.963 to 1.330, P = 0.044). PCKS9i significantly increased genetic risk of knee OA (OR = 1.136, 95% CI: 1.027 to 1.228, P = 0.013), but not for hip OA. Genetic risk of seropositive RA was strongly reduced while consuming PCSK9i (OR = 0.796, 95% CI: 0.580 to 0.964, P=0.020) and this effect is independent with LDL levels, while we don’t observe causal relationships with PCSK9i on seronegative RA.

**Conclusions:** This study elucidates the causal relationship between PCSK9i and genetic predisposition to OP, OA, and RA. PCSK9i would benefit LS-BMD and protect the genetic risk of seropositive RA. Meanwhile, PCSK9i might be a risk factor for knee OA.

## 1 Introduction

Hyperlipidemia, a well-established risk factor for cardiovascular diseases(1), has been increasingly implicated in the pathophysiology of bone disorders(2, 3). Osteoporosis (OP), osteoarthritis (OA), and rheumatoid arthritis (RA) are among the most prevalent bone diseases, each characterized by distinct pathological mechanisms yet sharing potential links with lipid metabolism(4, 5, 6).

Proprotein convertase subtilisin/kexin 9 inhibitors (PCSK9i), which could inhibit the PCSK9 protein to increase clearance of low-density lipoprotein cholesterol (LDL-C) from the bloodstream(7), have primarily been recognized for their role in hyperlipidemia treatment(8). However, emerging evidence suggests that PCSK9i has implications beyond lipid metabolism(9). In addition, another well-known cholesterol-lowering drug, statins have been reported to improve BMD, as well as reduce risk of fractures(10). This study aims to explore the potential impact of PCSK9i on bone diseases, in comparison to traditional statins, using Mendelian randomization (MR) to elucidate potential causal relationships.

While studies have started to unveil the relationship between hyperlipidemia and bone mineral density (BMD), the role of PCSK9i in this context remains less understood. The current research employs a comprehensive approach, utilizing single-nucleotide polymorphisms (SNPs) extracted from genome-wide association studies (GWAS) to investigate the influence of PCSK9i on OP, OA, and RA. This approach provides a robust framework to assess the impact of PCSK9i on these bone diseases, thereby contributing to a more nuanced understanding of the systemic effects of lipid-lowering agents. This study not only aims to clarify the role of PCSK9i in bone disease management but also seeks to offer insights into the broader implications of cholesterol metabolism in skeletal health.

## 2 Materials and Methods

### Data Acquisition and Processing

The BMD outcomes considered in this research pertained to BMD variation by specific skeletal sites. OA outcomes were split by hip and knee, and RA was characterized by seropositive or seronegative. Our foundational dataset incorporated GWAS summary outcomes: eBMD (N = 265,627), Knee-OA(n= 403,124), seronegative RA (n= 174,771) and seronegative RA (n= 177,430) from MRC-IEU and TB-BMD (N = 56,284), LS-BMD (N = 28,498), FN-BMD (N = 32,735), and FA-BMD (N = 8,143) from GEFOS(11), Hip-OA(n=14,275) from arcOGEN(12). Table 1 offers an overview of the data sources used and their respective participant demographics. The majority of participants were from European backgrounds. Given the study’s dependence on existing GWAS summary datasets, there was no need for institutional board approval, though consent was acquired from all contributors.

### Genetic instrument selection for PCSK9 inhibitors

Genetic associations with LDL cholesterol (LDL-C) was sourced from GWAS summary statistics encompassing 173,082 individuals(13). By identifying instrumental variables targeting PCSK9 to lower LDL-C, we aimed to mimic the effects of PCSK9 inhibitors(14). To circumvent linkage disequilibrium issues, we established a linkage disequilibrium threshold (r^2^ ≤ 0.3) and excluded SNPs that exhibited strong LD with each other. Following this process, 8 significant PCSK9i SNPs were retained, deemed appropriate as instrumental variables. In a complementary approach, FCHL as the positive control outcome, guided by the Consensus criteria (N = 39,961 cases; 309,261 controls) derived from UK Biobank(15).

### MR analysis

In our article, we highlight the importance of genetic variation as an instrumental variable (IV) in Mendelian Randomization (MR) studies. This requires adherence to three key assumptions: First, a significant correlation between the genetic variation and the exposure (e.g., protein levels); second, independence from confounding factors affecting both exposure and outcome; and third, the influence of the genetic variation on the outcome should occur solely through the exposure. Meeting these criteria ensures the accuracy and validity of causal inferences in MR studies.

The overarching objective of our Mendelian Randomization (MR) examination was to validate PCSK9i instrumental variables, as well as PCSK9i on BMD, OP, OA, and RA. Our primary modus operandi was the inverse variance weighted (IVW) technique(16). In a quest for heightened analytical rigor, we also engaged the MR-Egger regression and median-based estimator techniques(17). Situations with discernible horizontal pleiotropy saw the deployment of the MR-PRESSO outlier test(18). Furthermore, our analyses leveraged both IVW and MR Egger regression to probe horizontal pleiotropic SNP effects, with Cochran’s Q-test being instrumental(19) in heterogeneity quantifications. To extend our results, we performed the same analytical procedure for LDL cholesterol and bone disease.

### Robust analysis

We employed the Cochran Q-test metrics to measure heterogeneities. To evaluate possible horizontal pleiotropic impacts of the SNPs, we used IVW (random effect) and MR-Egger regression techniques. Moreover, we executed a “leave-one-out” sensitivity assessment to pinpoint any SNPs that might have a pronounced influence. In this approach, we sequentially removed each SNP to ascertain if it drove the observed association.

### Statistical Analysis

Bonferroni correction (corrected p = 0.05/X*Y, where X represents the number of exposures and Y represents the number of outcomes) was used to adjust for multiple tests in this MR analysis. All statistical analyses employed the Two-Sample MR (version 0.5.7) (20) R software (version 4.1.3).

## 3 Results

### 3.1 Positive control analysis for PCSK9 inhibitors instrumental variables

PCSK9 inhibitors (PCSK9i), known for their effectiveness in hyperlipidemia treatment. Using the ieu-a300 dataset, we identified eight significant SNPs associated with PCSK9 inhibitors as instrumental variables (Table 2). To validate identified PCSK9i instrumental variables, we chose GWAS summary data of Familial Combined Hyperlipidemia (FCHL), which is characterized by high blood cholesterols(21) as positive controls for PCSK9i instrumental variables. As expected, our Mendelian Randomization (MR) analysis revealed that PCSK9i significantly reduced the risk of FCHL (Odds Ratio: 0.3311, 95% CI: 0.286-0.383, P.adj = 3.2e-50; Figure 1).

**Figure 1.**
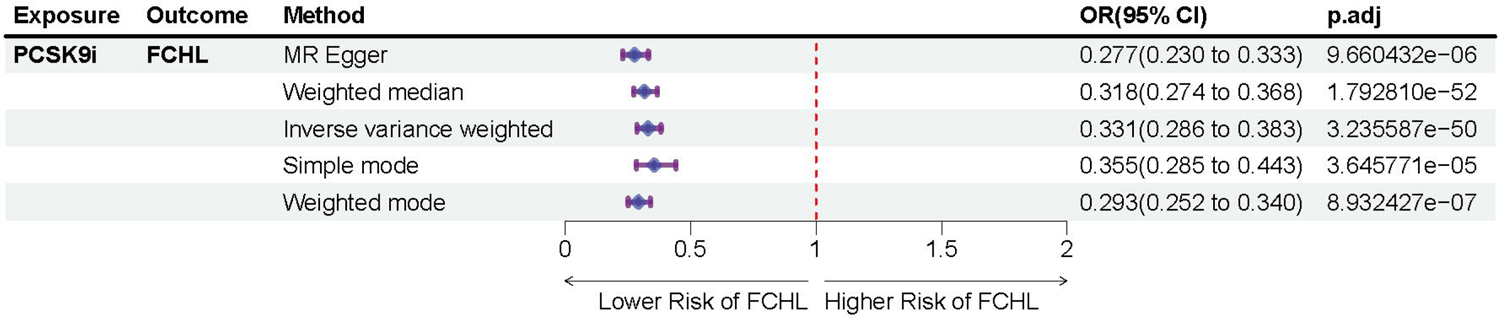
Causal impact of PCSK9i on FCHL. The Forest plot was drawn using IVW results. OR: odds ratio; IVW: inverse variance weighting; CI: confidence interval.

### 3.2 PCSK9 inhibitor usage associated with higher lumbar spine BMD

We next conduct two sample Mendelian Randomization analysis for PCSK9i and osteoporosis (OP), which is marked by reduced Bone Mineral Density (BMD) and impaired bone integrity(22). In conclusion, our results have not shown a causal effect between PCSK9i on susceptibility of OP neither postmenopausal osteoporosis (PMOP) with fracture. However, a positive association of PCSK9 inhibitors with increased Lumbar Spine-BMD (LS-BMD; OR=1.157, 95% CI: 0.963 - 1.330, p.adj=0.0442) was identified (Figure 2). The relationship we did is similar with previous research(23), as shown in Supplementary Figure 1.

**Figure 2.**
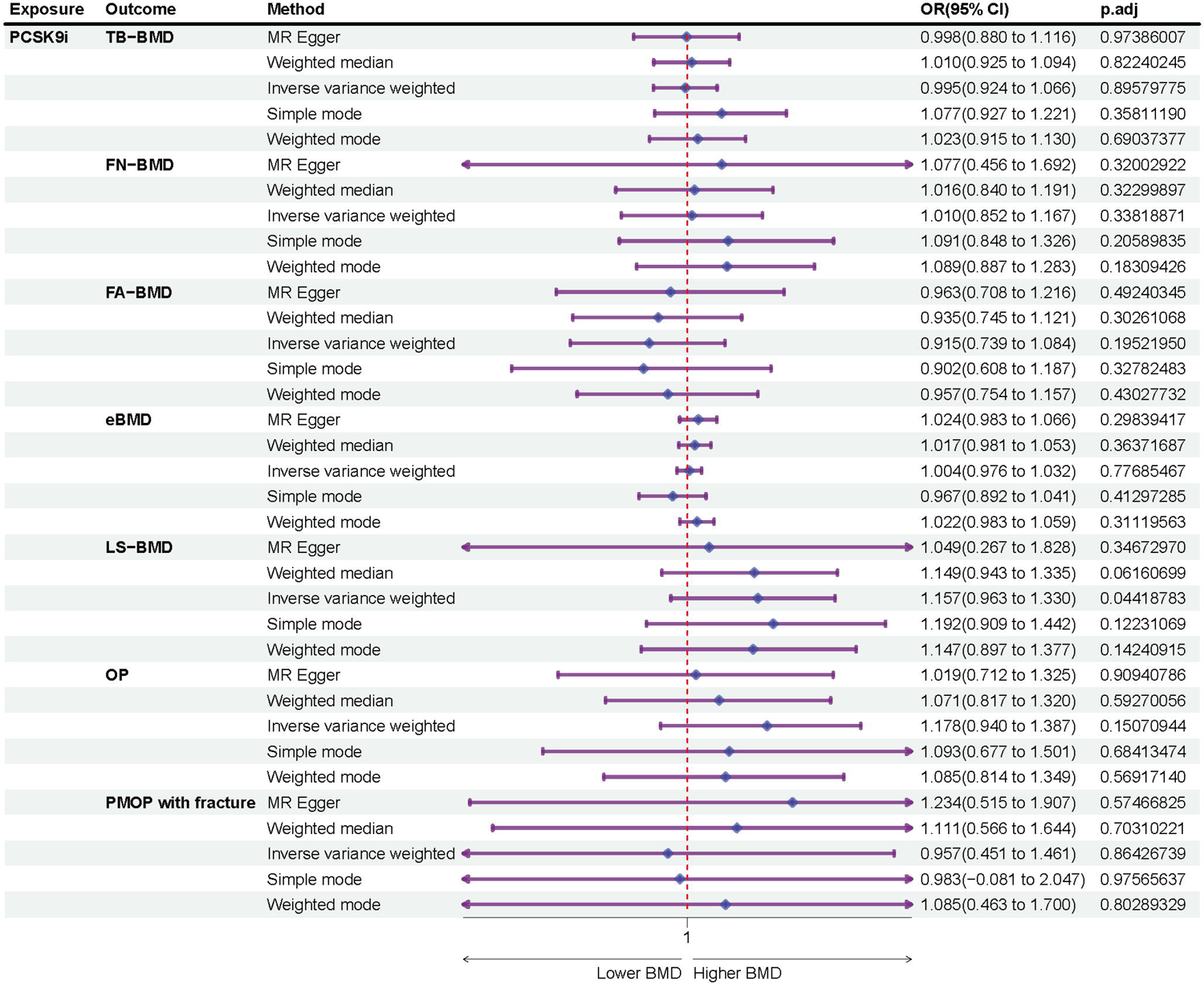
Causal impact of PCSK9i on OP and BMD. The Forest plot was drawn using IVW results. OR: odds ratio; IVW: inverse variance weighting; CI: confidence interval.

### 3.3 PCSK9i intake leads to high genetic risk of knee OA

We utilized data from two Genome-Wide Association Studies (GWAS) cohorts diagnosed with hip and knee osteoarthritis, respectively. This approach allowed us to explore the potential influence of PCSK9i on these specific subtypes of osteoarthritis at a genetic level.

While we found no causal relationship between PCSK9i intake and hip osteoarthritis, we observed a different scenario for knee osteoarthritis. Notably, our results indicated that the genetic risk of knee osteoarthritis was significantly increased with PCSK9i intake (OR=1.136, 95% CI: 1.027-1.228, p.adj=0.0126, Figure 3). MR analysis of LDL and genetic risk for knee OA suggests that this effect is related to reduced LDL levels (OR=0.909, 95% CI: 0.859-0.951, p.adj=5.46e-5, Figure 3).

**Figure 3.**
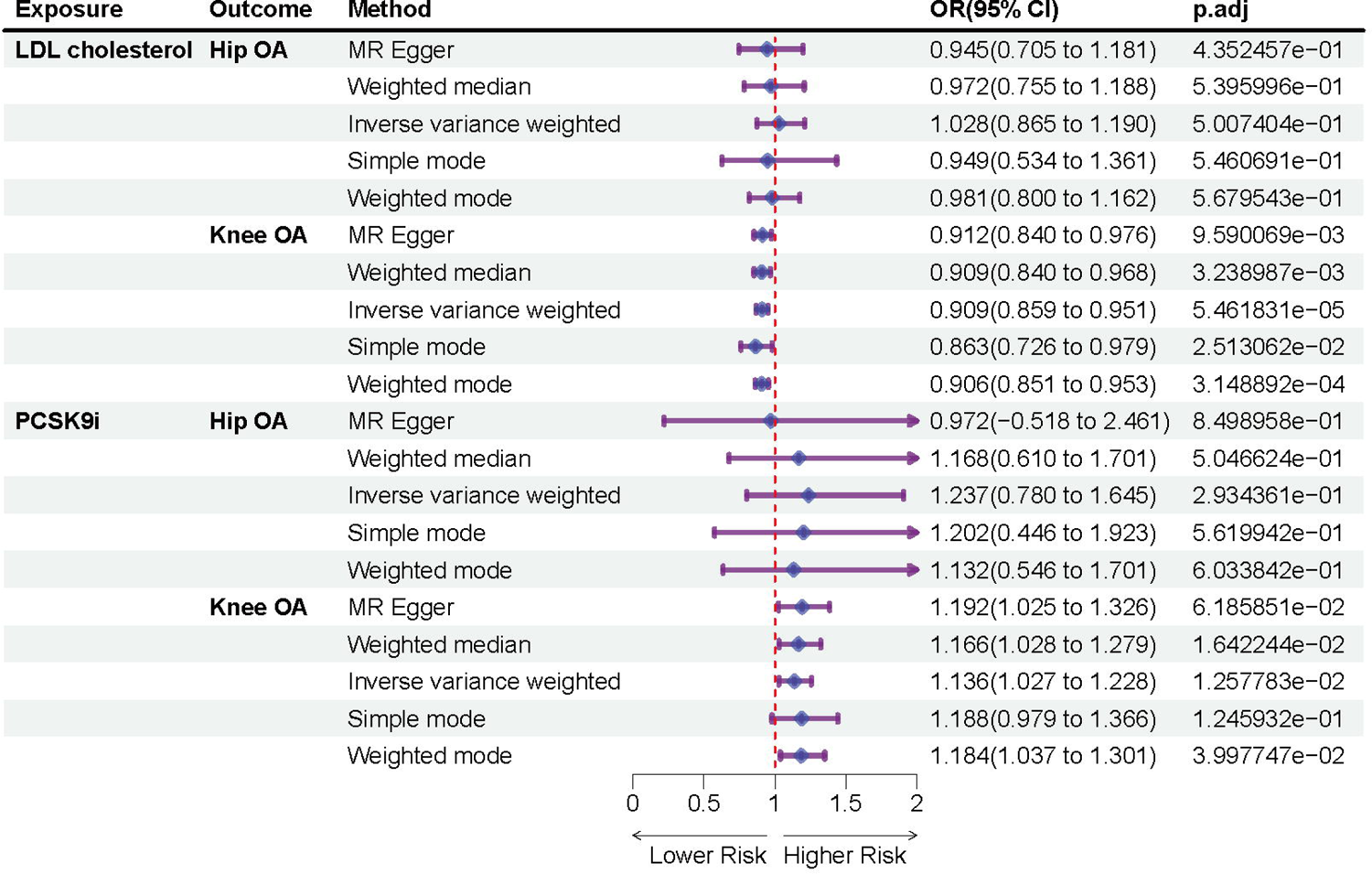
Causal impact of PCSK9i on hip OA and knee OA. The Forest plot was drawn using IVW results. OR: odds ratio; IVW: inverse variance weighting; CI: confidence interval.

### 3.4 PCSK9i intake reduces the risk of seropositive RA and is independent of LDL levels

Rheumatoid arthritis (RA) is a chronic inflammatory disease that affects the joints. RA can be classified into seropositive RA and seronegative RA based on the presence or absence of rheumatoid factor (RF) and anti-citrullinated protein antibodies (ACPA) in the blood(24). Seropositive RA is more common and tends to have a more severe course and worse prognosis than seronegative RA(25, 26). A notable outcome of our study is the lack of evidence supporting a significant risk association between PCSK9i and seronegative RA. More interestingly, the analysis revealed that the genetic susceptibility to seropositive RA could be significantly mitigated by the intake of PCSK9i (OR=0.796, 95% CI: 0.580-0.964, p.adj=0.0202, Figure 4). The subsequent LDL MR analysis corroborates this conclusion, showing that the beneficial role of PCSK9i in seropositive RA is not a consequence of LDL reduction, since LDL levels do not correlate with genetic risk of seropositive RA (OR=1.034, 95% CI: 0.932-1.135, p.adj=0.506, Figure 4). This observation is particularly significant as it implies a potential therapeutic role of PCSK9i in the management of seropositive RA, independent of its lipid-lowering effects.

**Figure 4.**
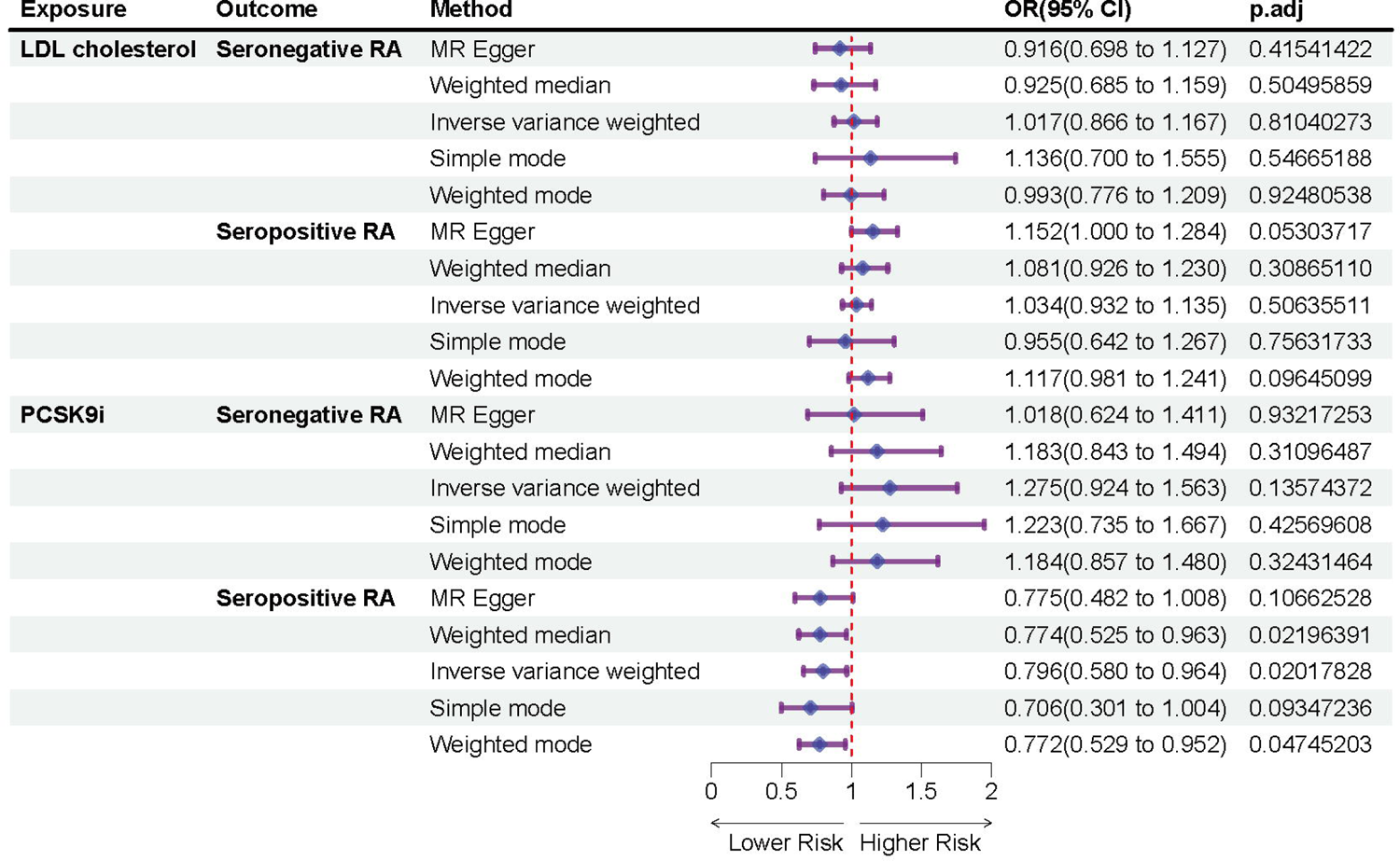
Causal impact of PCSK9i on seronegative RA and seropositive RA. The Forest plot was drawn using IVW results. OR: odds ratio; IVW: inverse variance weighting; CI: confidence interval.

### 3.5 Robustness

The MR-Egger regression method was utilized to investigate the potential existence of horizontal pleiotropy among the SNPs and the result. Our results did not identify any evidence of such pleiotropy (all p > 0.05) (Supplementary Tables 1). Similarly, the funnel plots did not display any apparent horizontal pleiotropy for the evaluated outcomes (Supplementary Figure 2). Additionally, the leave-one-out sensitivity charts revealed that no individual SNP significantly impacted the causal association, emphasizing the strength of our conclusions (Supplementary Figure 3).

## 4 Discussion

Although previous studies indicating that a reduction in genetically predicted low-density lipoprotein cholesterol (LDL-C) is linked to an increase in bone mineral density(23), furthermore the osteoporosis(27). Our finding that PCSK9i does not significantly impact overall osteoporosis (OP) risk contrasts with some literature suggesting statins may positively influence bone health(28). This discrepancy underscores the complexity of lipid-lowering agents’ effects on bone metabolism. While statins are thought to exert osteoanabolic effects possibly via the mevalonate pathway(29), the mechanism through which PCSK9i might affect bone density remains less clear. The observed increase in lumbar-spine bone mineral density (LS-BMD) with PCSK9i use, demands a more refined understanding, particularly when contrasted with the broader systemic effects of statins.

The study’s revelation of an increased genetic risk for knee osteoarthritis (OA) but not hip OA. This specificity suggests a potential biomechanical or local metabolic factor influenced by PCSK9i that differentially affects joints. The mechanistic pathways linking cholesterol metabolism, PCSK9i action, and joint-specific OA development need to be rigorously investigated. This could pave the way for joint-specific therapeutic strategies and possibly uncover unforeseen side effects of PCSK9i therapy in susceptible individuals. Statins have been hypothesized to exert anti-inflammatory and chondroprotective effects(30), potentially beneficial in OA management(31). The joint-specific risk increase associated with PCSK9i raises critical questions about the differential impact of cholesterol-lowering drugs on joint health. It suggests that the systemic lipid-lowering effect might not uniformly translate into protective benefits across all joints.

The discovery that PCSK9 inhibitors can reduce the genetic risk of seropositive rheumatoid arthritis (RA) is a significant advancement in understanding the interplay between lipid metabolism and autoimmune diseases. However, an intriguing aspect of this finding is the lack of a direct correlation between low-density lipoprotein (LDL) cholesterol levels and the risk of RA. This suggests that the beneficial effects of PCSK9 inhibitors in reducing the risk of seropositive RA might not be solely due to their primary role in lowering LDL cholesterol levels.

One possible explanation for this phenomenon could be the pleiotropic effects of PCSK9 inhibitors. Beyond their well-known impact on lipid levels, PCSK9 inhibitors may exert additional biological effects that influence immune response and inflammation(32, 33), two key factors in the pathogenesis of RA. Another hypothesis is that the mechanism by which PCSK9 inhibitors reduce RA risk may involve modulation of lipid subfractions or other metabolic pathways(34), rather than a straightforward reduction in LDL cholesterol. Alterations in these components can have significant effects on inflammatory processes and immune responses, potentially contributing to the observed reduction in RA risk.

Considering these findings, it is paramount to adopt a critically analytical lens towards the application of PCSK9i in bone diseases. While the potential benefits in LS-BMD enhancement and seropositive RA risk reduction are promising, the risk of exacerbating knee OA poses a significant clinical dilemma. Future research should aim to dissect the molecular and cellular pathways through which PCSK9i exerts these differential effects on bone and joint health. Investigating the role of PCSK9i in bone remodeling, joint-specific cartilage metabolism, and immune modulation could unravel the complex interplay between lipid metabolism and musculoskeletal diseases. Ultimately, these efforts could aid in developing more nuanced and effective treatment strategies for bone diseases.

## 5 Conclusion

This research clarifies the causal link between PCSK9 inhibitors (PCSK9i) and genetic susceptibility to osteoporosis (OP), osteoarthritis (OA), and rheumatoid arthritis (RA). The study suggests that PCSK9i positively impacts lumbar spine bone mineral density (LS-BMD) and may offer protective effects against the genetic risk associated with seropositive RA. However, it also indicates that PCSK9i could potentially be a risk factor for the development of knee OA.

## Data Availability

The datasets analyzed for this study can be found in the IEU OpenGWAS https://gwas.mrcieu.ac.uk/datasets/.

## Abbreviation

MR: Mendelian Randomization
IVs: Instrumental variables
FCHL: Familial combined hyperlipidemia
BMD: Bone mineral density
TB-BMD: Total body-bone mineral density
LS-BMD: Lumbar spine bone mineral density
FN-BMD: Femoral neck bone mineral density
FA-BMD: Forearm bone mineral density
eBMD: Heel bone mineral density
RA: Rheumatoid osteoarthritis
OP: Osteoporosis
PMOP: postmenopausal osteoporosis
OA: Osteoarthritis
IVW: Inverse variance weighted
SNPs: single nucleotide polymorphisms
OR: Odds ratio

## 7 Declarations

### 7.1 Ethics approval and consent to participate

Not applicable.

### 7.2 Consent for publication

Not applicable.

### 7.3 Competing interests

The authors declare that the research was conducted in the absence of any commercial or financial relationships that could be construed as a potential conflict of interest.

### 7.5 Funding

This work was supported by Natural Science Basic Research Program of Shaanxi(Program No.2025JC-YBQN-1050)

### 7.6 Authors’ contributions

Conceptualization, Dingjun Hao; Data curation, Xiaohui Wang; Formal analysis, Wangyu Yang; Funding acquisition, Xiaohui Wang; Supervision, Dingjun Hao; Visualization, Xiaohui Wang; Writing – original draft, Wangyu Yang; Writing – review & editing, Xiaohui Wang, Hao Yang and Dingjun Hao. This study was completed with teamwork. Each author had made corresponding contribution to the study. All authors contributed to the article and approved the submitted version.

## Notes

### Competing Interest Statement

The authors have declared no competing interest.

### Author Declarations

The study used ONLY openly available human data that were originally located at: https://gwas.mrcieu.ac.uk/

### Summary of Updates

The author affiliations have been updated to accurately reflect the current institutional positions and contributions of the authors at the time of manuscript submission. This revision ensures that institutional credits are correctly assigned and that correspondence and academic attribution align with the authors present affiliations. No other content of the manuscript has been altered.

