## Supplementary material for "Impact of PCSK9 inhibitors on bone disease: A comprehensive drug-target Mendelian randomization study": Supplementary Figure 1.pdf

| Outcome | Method | OR(95% CI) | p.adj |
|---|---|---|---|
| **FN−BMD** | MR Egger | 0.963(0.857 to 1.068) | 2.183963e−01 |
| | Weighted median | 0.983(0.887 to 1.080) | 3.255901e−01 |
| | Inverse variance weighted | 0.980(0.918 to 1.041) | 2.292896e−01 |
| | Simple mode | 0.962(0.806 to 1.117) | 2.788402e−01 |
| | Weighted mode | 0.976(0.888 to 1.063) | 2.595362e−01 |
| **FA−BMD** | MR Egger | 1.076(0.917 to 1.228) | 2.329374e−01 |
| | Weighted median | 1.036(0.900 to 1.171) | 3.886172e−01 |
| | Inverse variance weighted | 1.013(0.914 to 1.112) | 5.125168e−01 |
| | Simple mode | 0.950(0.678 to 1.219) | 4.552833e−01 |
| | Weighted mode | 1.036(0.900 to 1.170) | 3.936511e−01 |
| **eBMD** | MR Egger | 0.968(0.942 to 0.992) | 1.214070e−02 |
| | Weighted median | 0.974(0.955 to 0.992) | 3.965679e−03 |
| | Inverse variance weighted | 0.970(0.952 to 0.987) | 6.073608e−04 |
| | Simple mode | 0.953(0.917 to 0.987) | 8.697523e−03 |
| | Weighted mode | 0.970(0.955 to 0.983) | 5.854258e−05 |
| **LS−BMD** | MR Egger | 1.074(0.937 to 1.206) | 1.361648e−01 |
| | Weighted median | 1.071(0.961 to 1.176) | 9.475364e−02 |
| | Inverse variance weighted | 0.965(0.883 to 1.045) | 1.706505e−01 |
| | Simple mode | 0.984(0.763 to 1.206) | 3.957068e−01 |
| | Weighted mode | 1.038(0.939 to 1.136) | 2.032840e−01 |
| **OP** | MR Egger | 1.034(0.870 to 1.198) | 6.700755e−01 |
| | Weighted median | 0.945(0.771 to 1.115) | 5.050821e−01 |
| | Inverse variance weighted | 1.007(0.893 to 1.120) | 8.854043e−01 |
| | Simple mode | 0.997(0.671 to 1.323) | 9.616756e−01 |
| | Weighted mode | 0.975(0.807 to 1.142) | 7.478573e−01 |
| **PMOP with fracture** | MR Egger | 0.904(0.535 to 1.263) | 5.742119e−01 |
| | Weighted median | 0.883(0.478 to 1.273) | 5.254661e−01 |
| | Inverse variance weighted | 1.067(0.811 to 1.319) | 6.008979e−01 |
| | Simple mode | 0.882(0.127 to 1.622) | 7.246108e−01 |
| | Weighted mode | 0.862(0.480 to 1.223) | 4.252994e−01 |
| **TB−BMD** | MR Egger | 0.981(0.937 to 1.024) | 3.777300e−01 |
| | Weighted median | 0.981(0.939 to 1.023) | 3.707564e−01 |
| | Inverse variance weighted | 0.962(0.931 to 0.991) | 9.915787e−03 |
| | Simple mode | 1.012(0.922 to 1.102) | 7.817796e−01 |
| | Weighted mode | 0.985(0.946 to 1.023) | 4.233605e−01 |

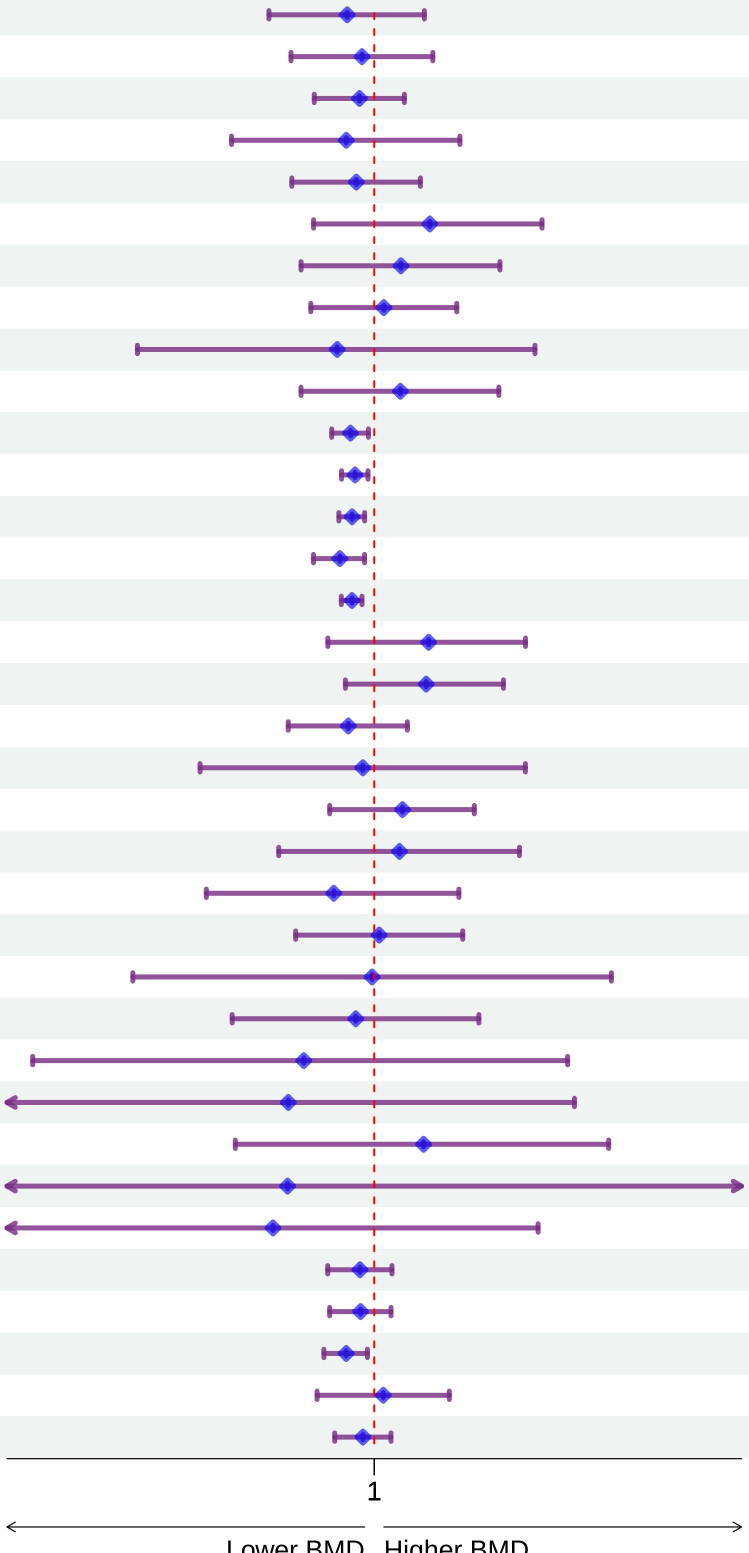
