## Supplementary material for "Impact of PCSK9 inhibitors on bone disease: A comprehensive drug-target Mendelian randomization study": Supplementary Figure 2.pdf

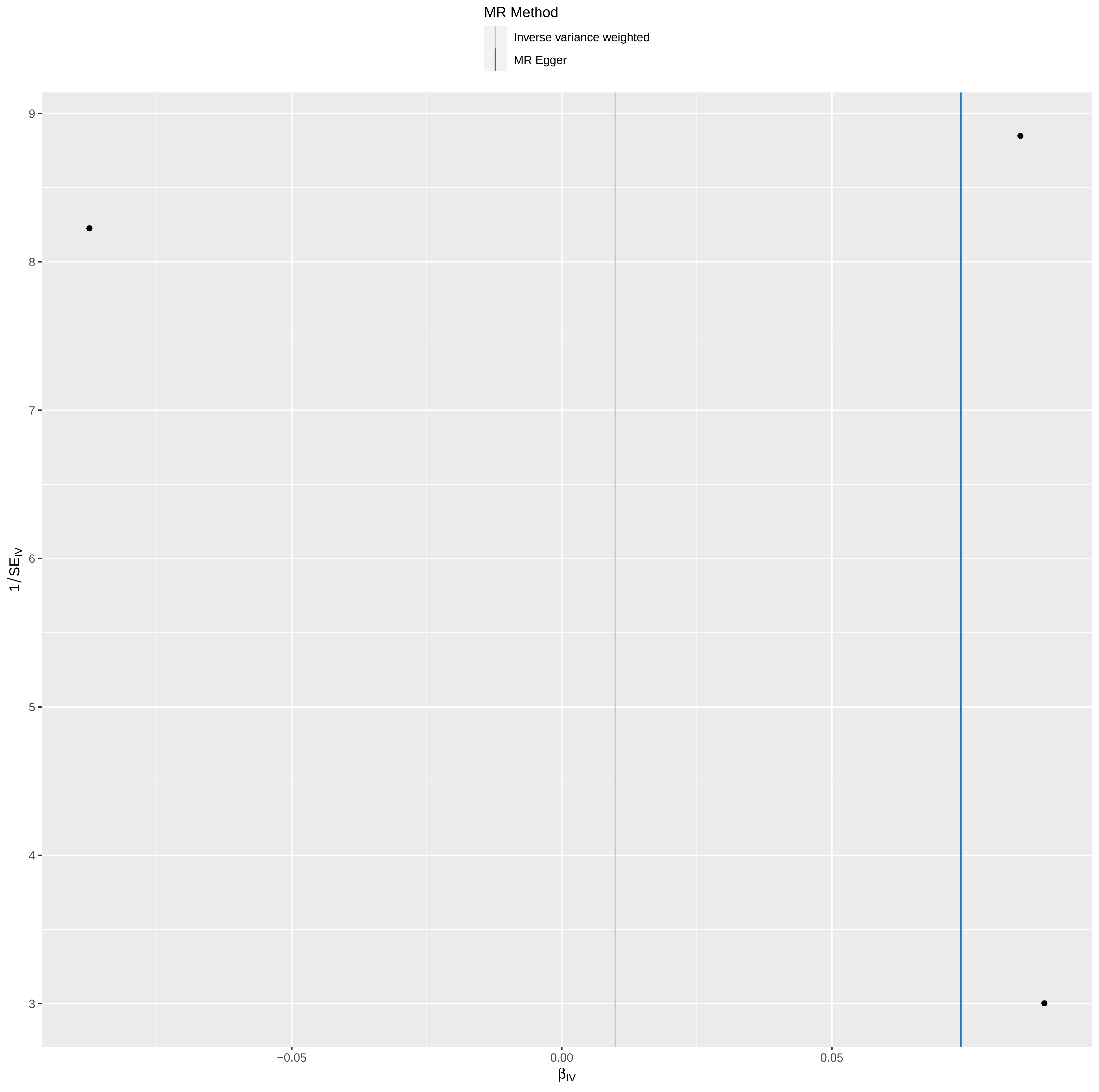

MR Method
Inverse variance weighted
MR Egger
9
8
7
6
5
4
3
1/SE_IV
−0.05
0.00
0.05
β_IV

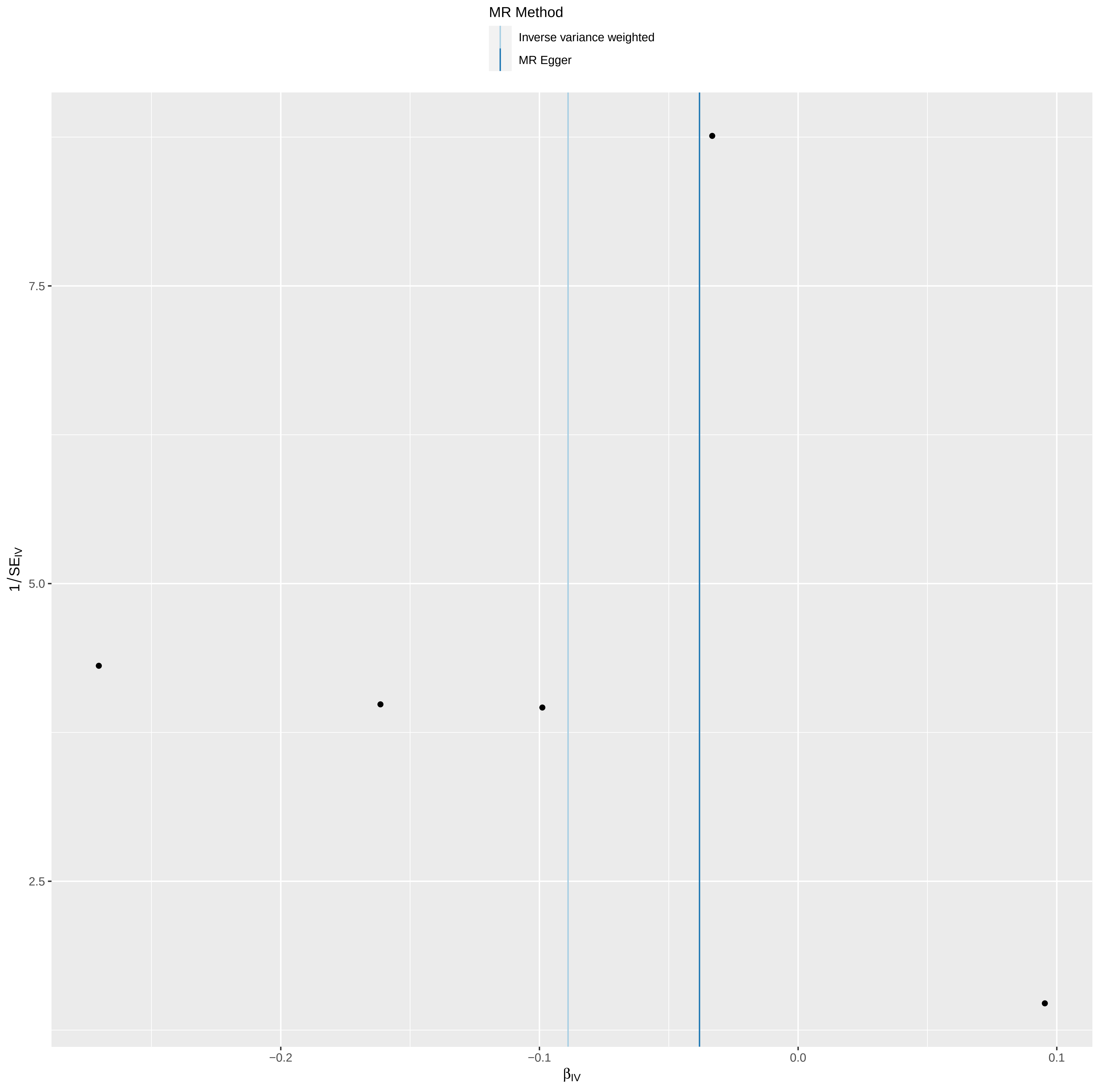
MR Method
Inverse variance weighted
MR Egger
7.5
5.0
2.5
1/SE_IV
−0.2
−0.1
0.0
0.1
β_IV

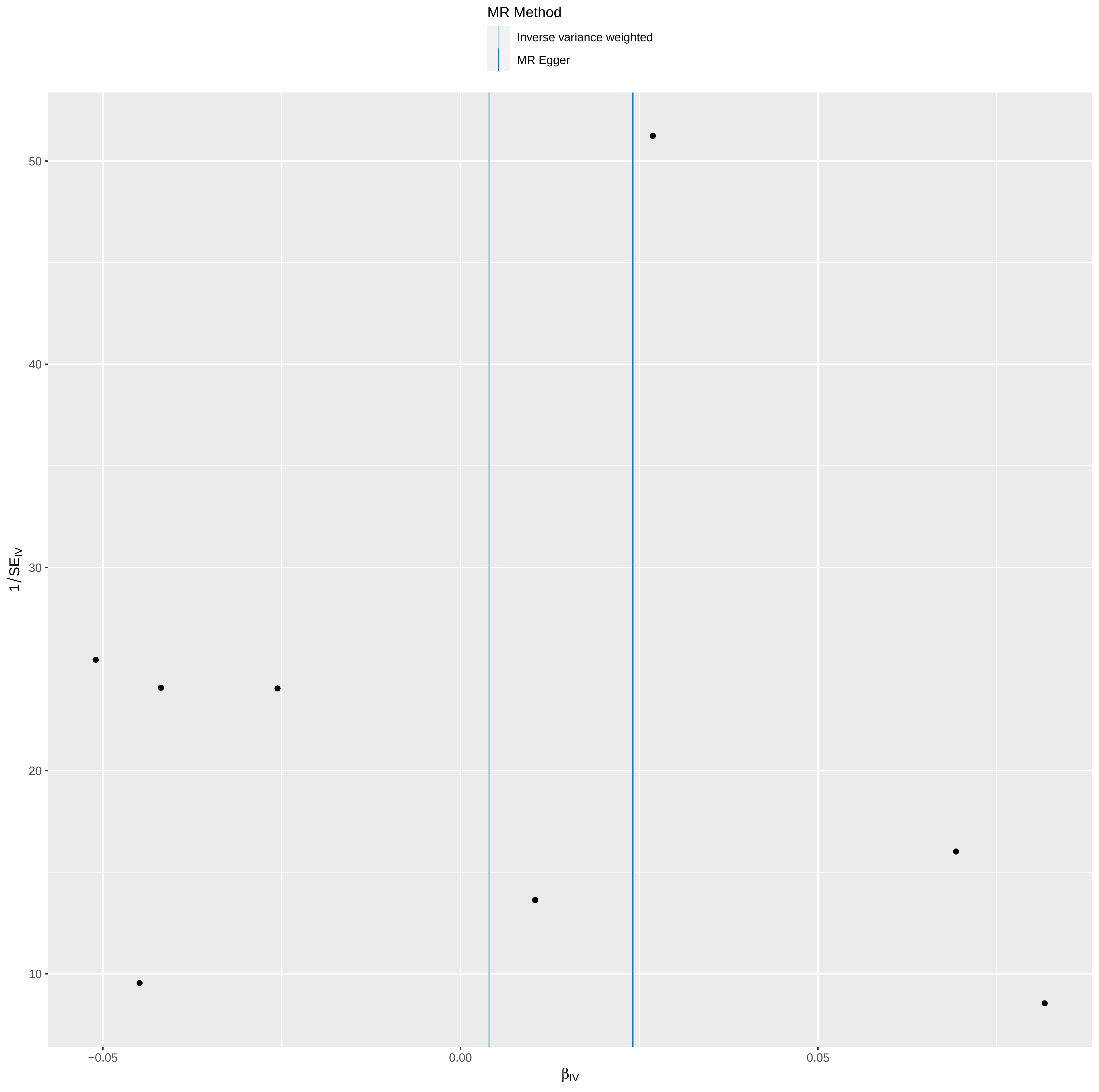
MR Method
Inverse variance weighted
MR Egger
50
40
30
20
10
$1/SE_{IV}$
−0.05
0.00
0.05
$\beta_{IV}$

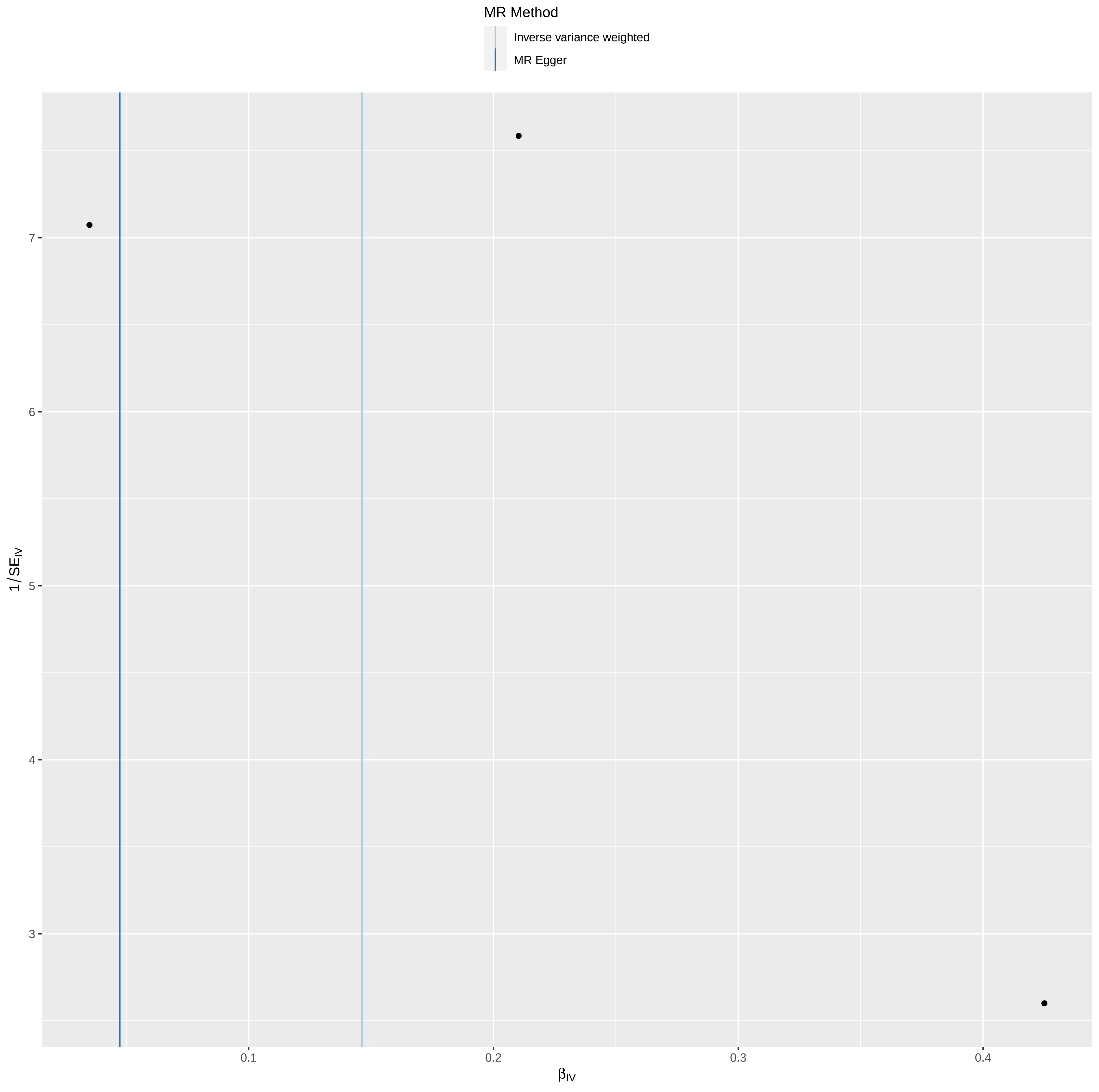

MR Method
Inverse variance weighted
MR Egger
1/SE_IV
7
6
5
4
3
0.1
0.2
0.3
0.4
β_IV

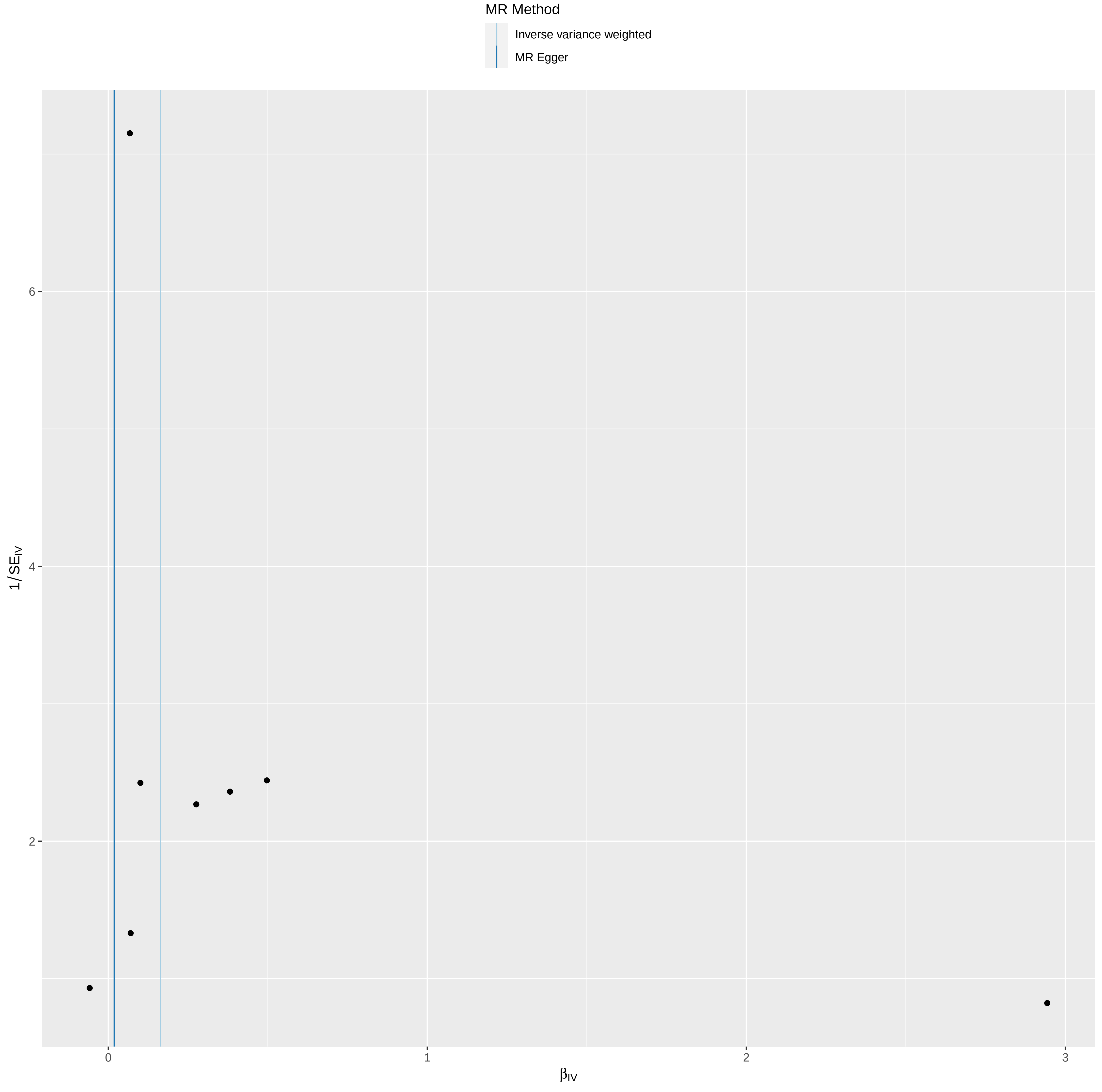
MR Method
Inverse variance weighted
MR Egger
1/$SE_{IV}$
6
4
2
0
1
2
3
$\beta_{IV}$

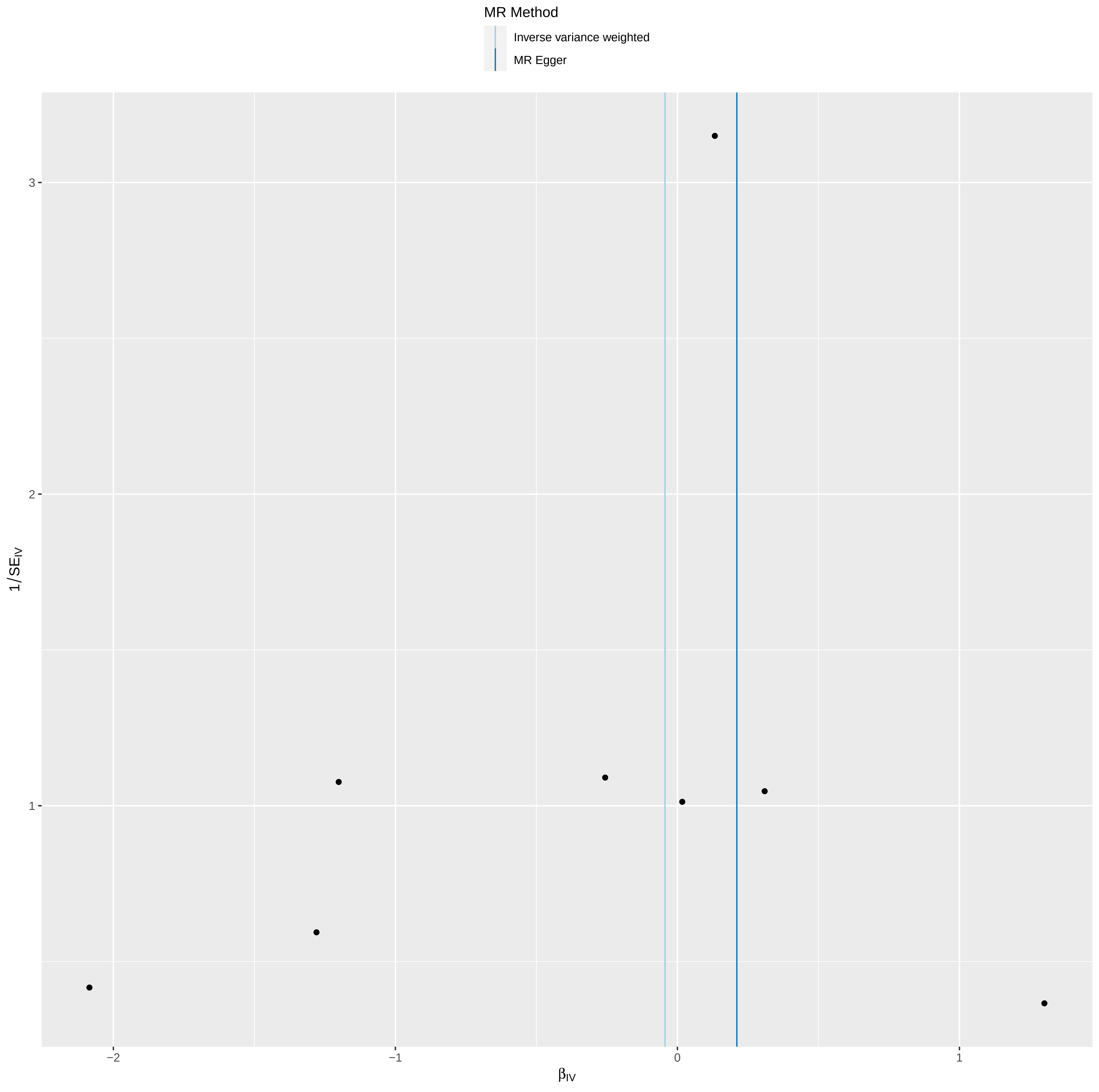

MR Method
Inverse variance weighted
MR Egger
1/SE$_{IV}$
3
2
1
−2
−1
0
1
$\beta_{IV}$

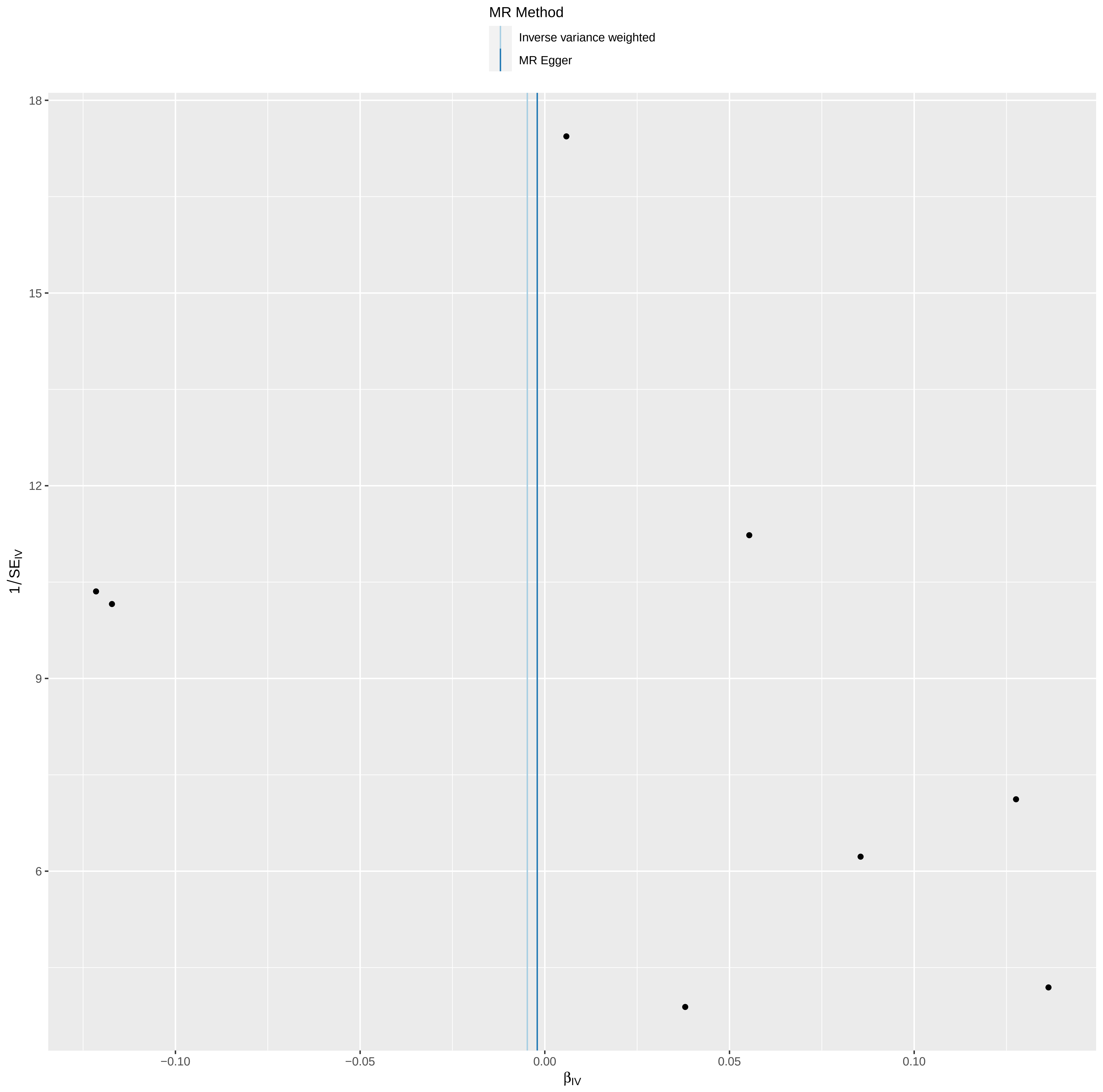

MR Method
Inverse variance weighted
MR Egger
18
15
12
9
6
1/SE_IV
−0.10
−0.05
0.00
0.05
0.10
β_IV

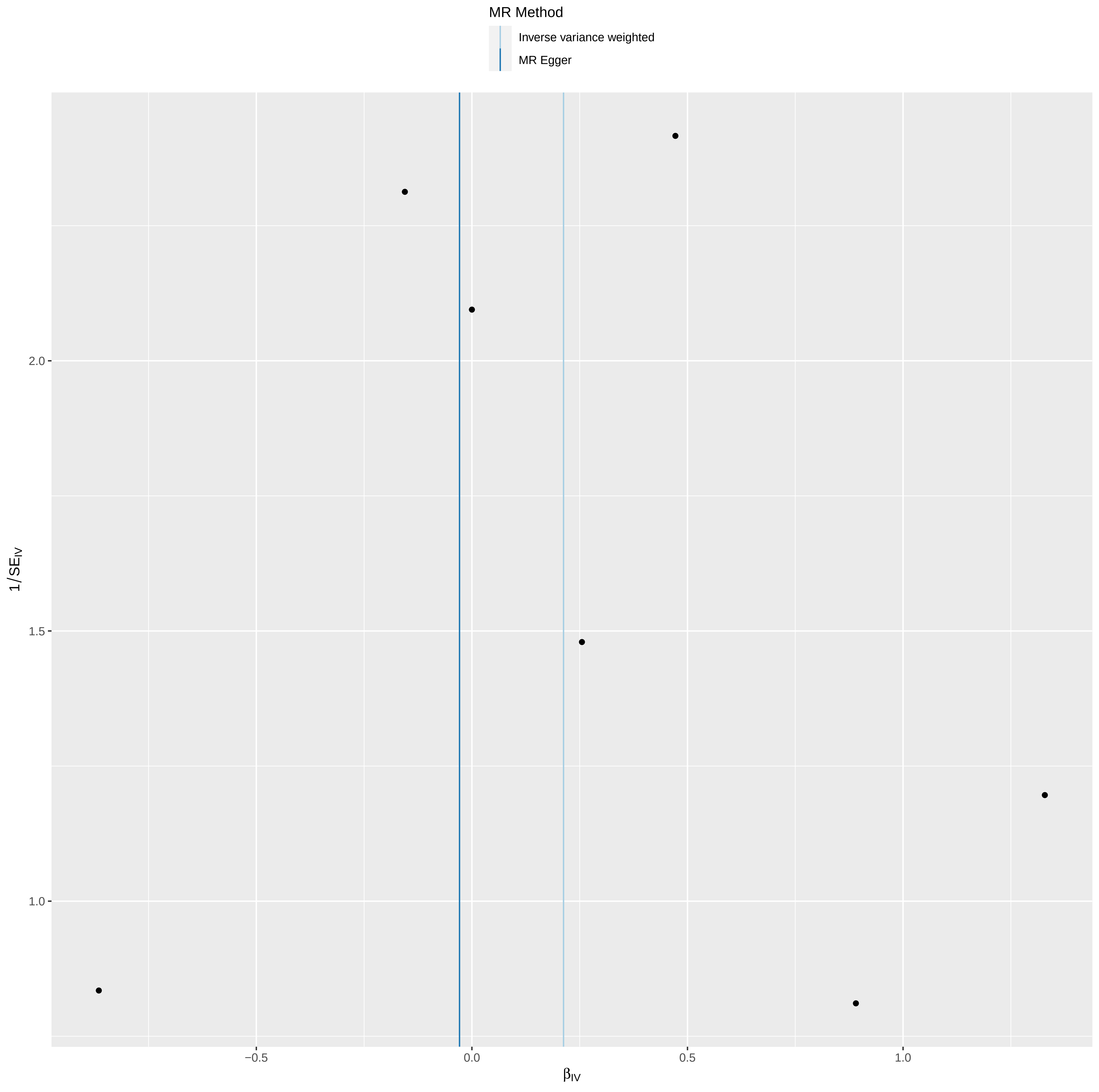

MR Method
Inverse variance weighted
MR Egger
1/SE_IV
2.0
1.5
1.0
−0.5
0.0
0.5
1.0
β_IV

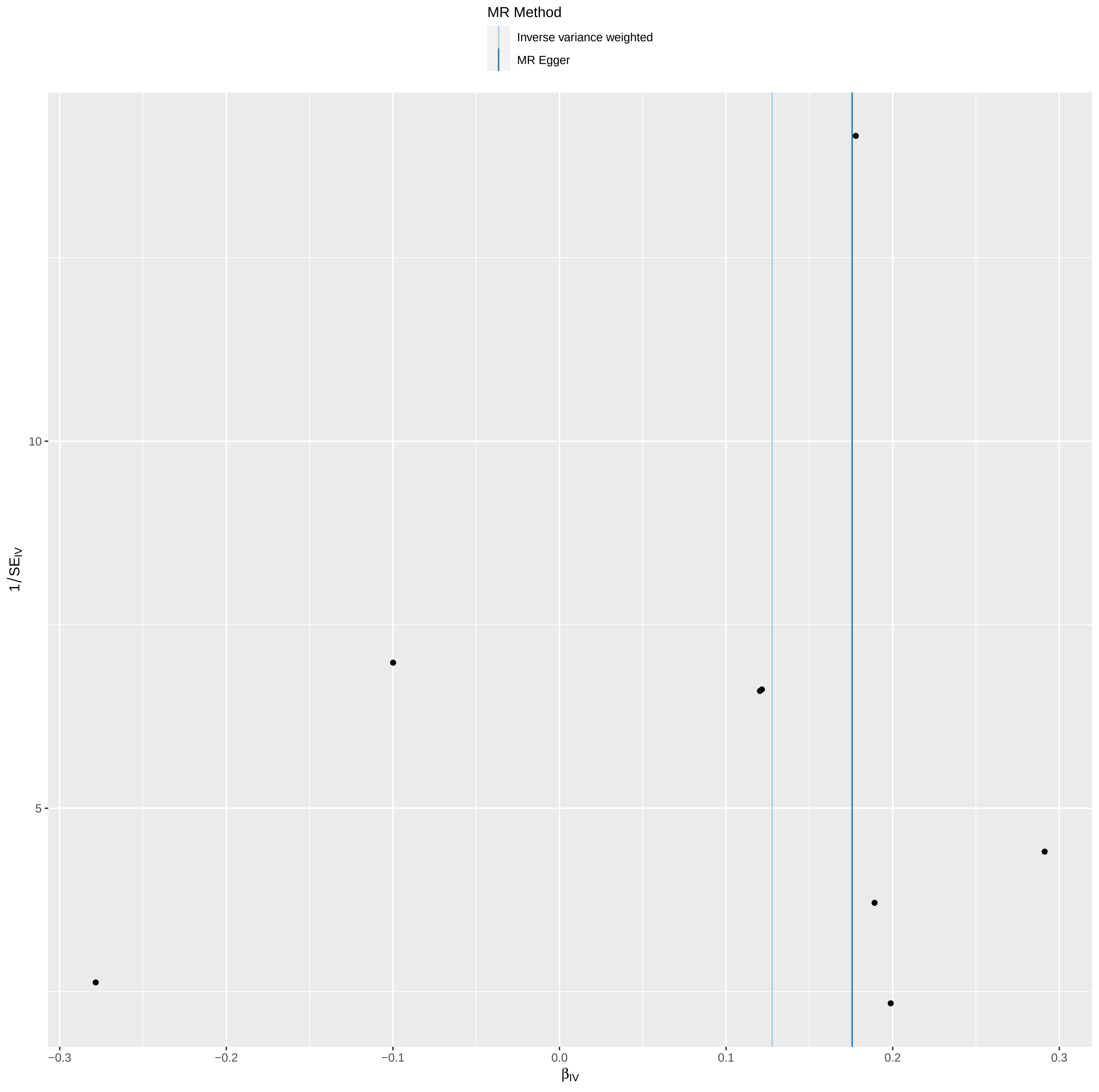
MR Method
Inverse variance weighted
MR Egger
1/SE_IV
10
5
−0.3
−0.2
−0.1
0.0
0.1
0.2
0.3
β_IV

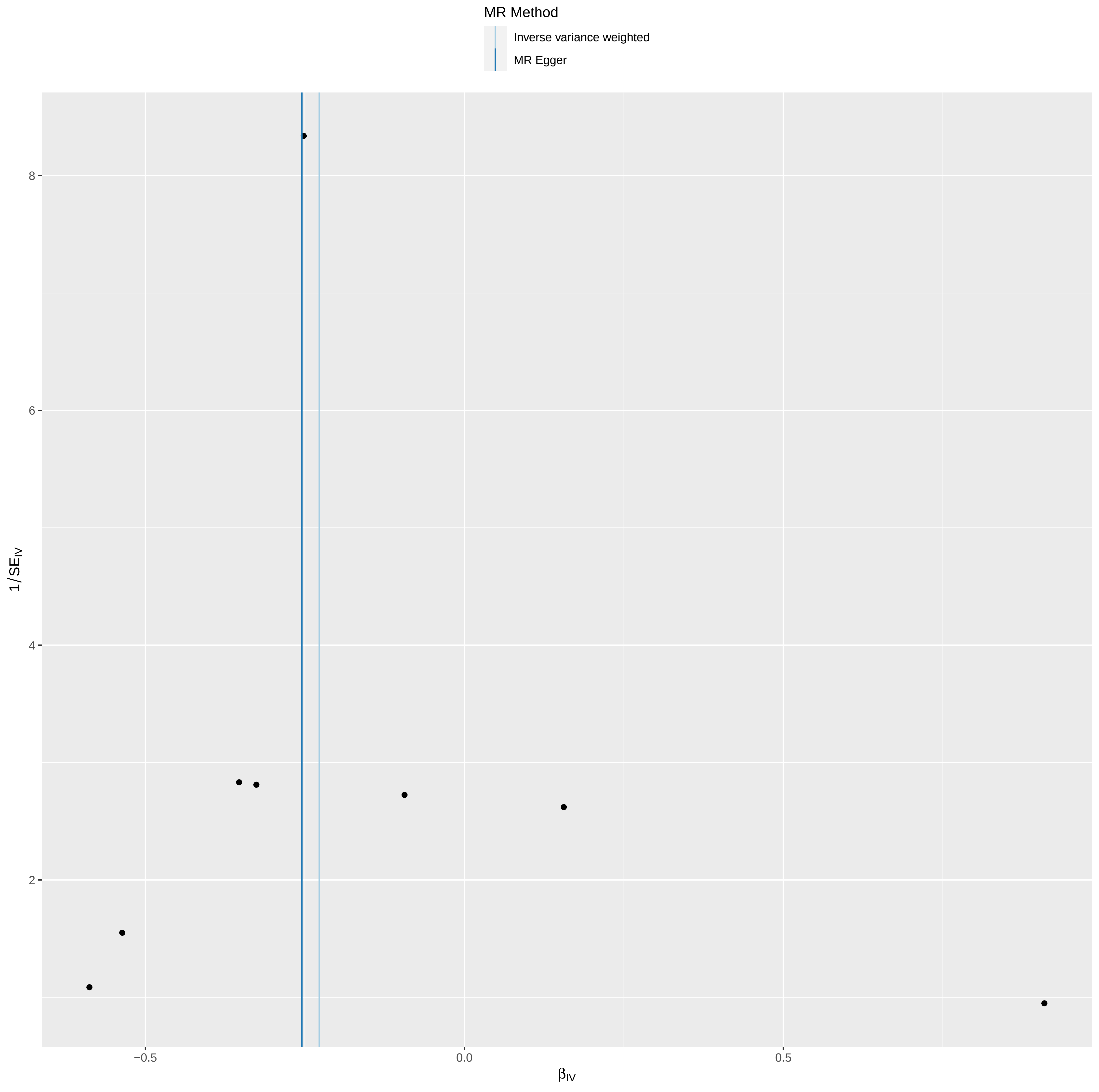

MR Method
Inverse variance weighted
MR Egger
8
6
4
2
1/SE$_{IV}$
−0.5
0.0
0.5
β$_{IV}$

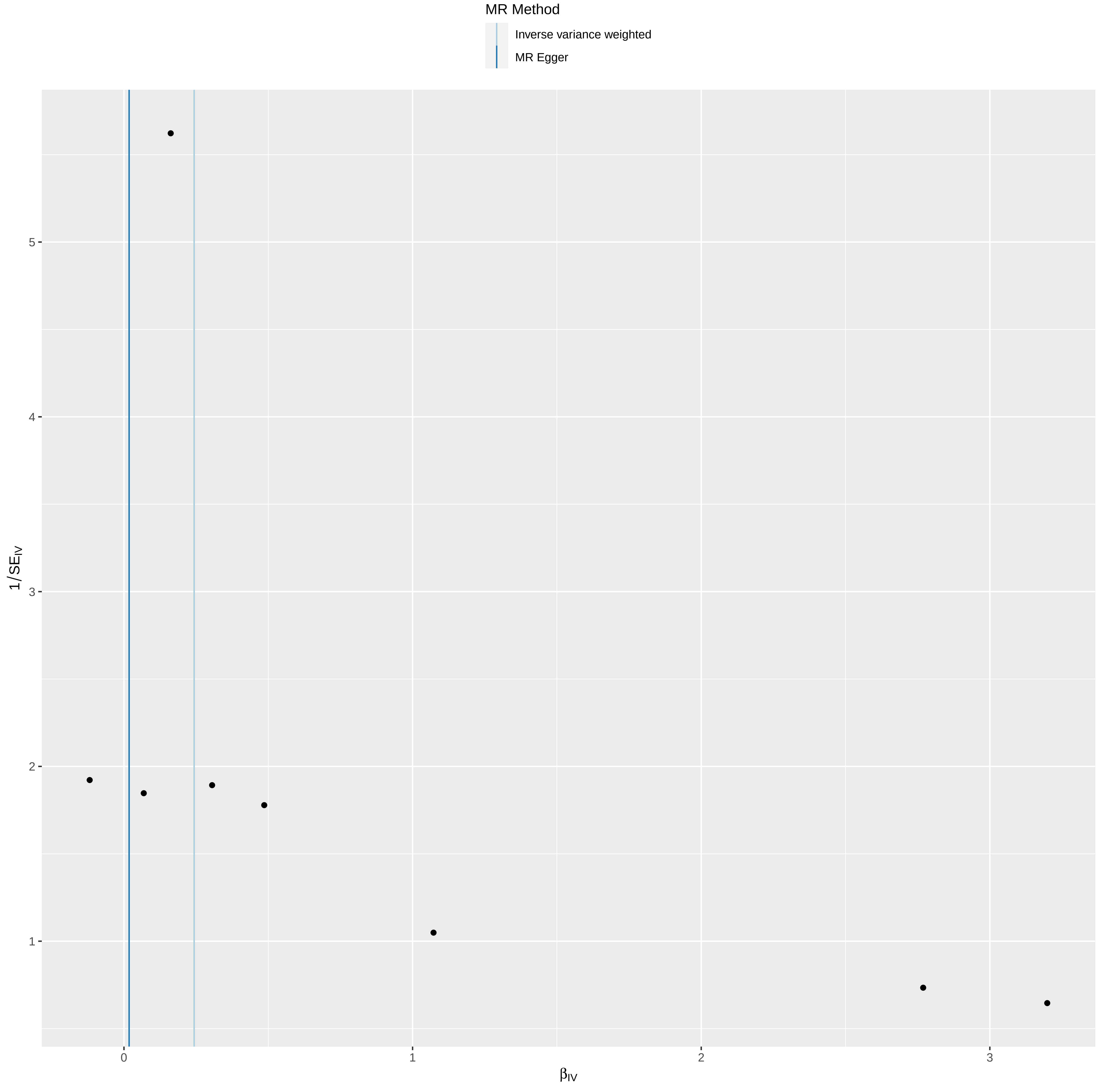

MR Method
Inverse variance weighted
MR Egger
1/SE_IV
5
4
3
2
1
0
1
2
3
β_IV
