## Supplementary material for "Impact of PCSK9 inhibitors on bone disease: A comprehensive drug-target Mendelian randomization study": Supplementary Figure 3.pdf

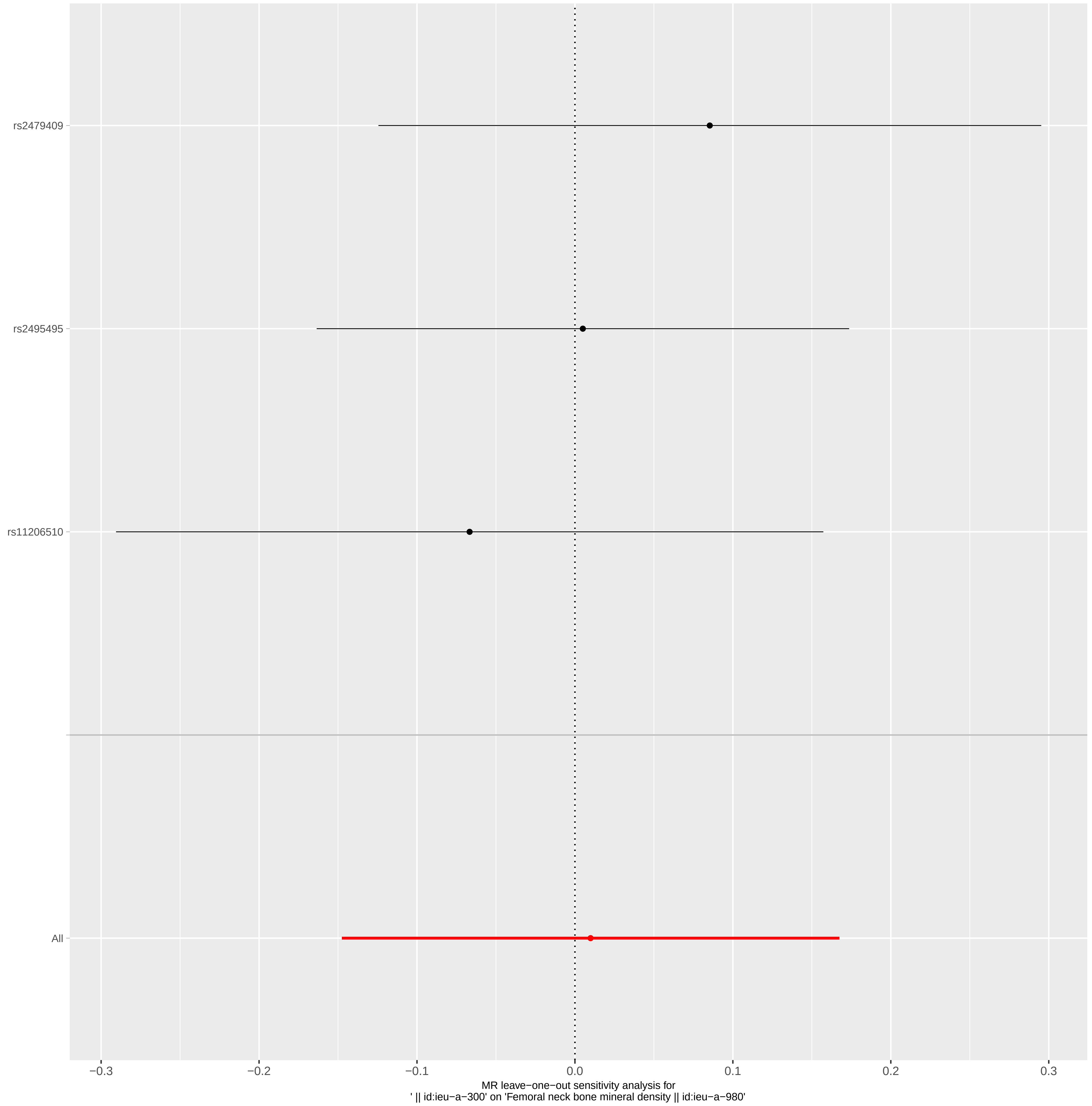

rs2479409
rs2495495
rs11206510
All
−0.3
−0.2
−0.1
0.0
0.1
0.2
0.3
MR leave−one−out sensitivity analysis for
' || id:ieu−a−300' on 'Femoral neck bone mineral density || id:ieu−a−980'

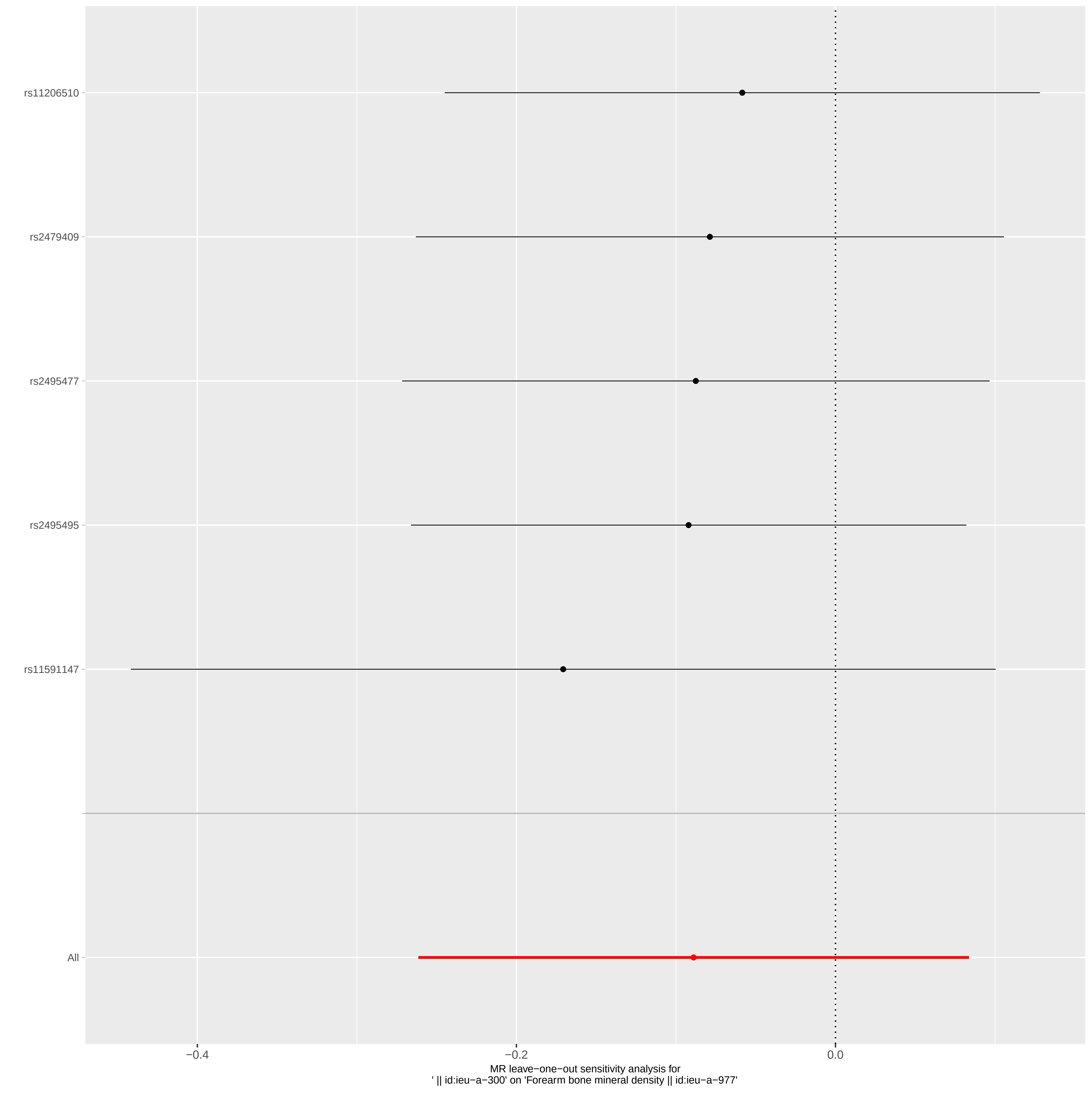
rs11206510
rs2479409
rs2495477
rs2495495
rs11591147
All
−0.4
−0.2
0.0
MR leave−one−out sensitivity analysis for
' || id:ieu−a−300' on 'Forearm bone mineral density || id:ieu−a−977'

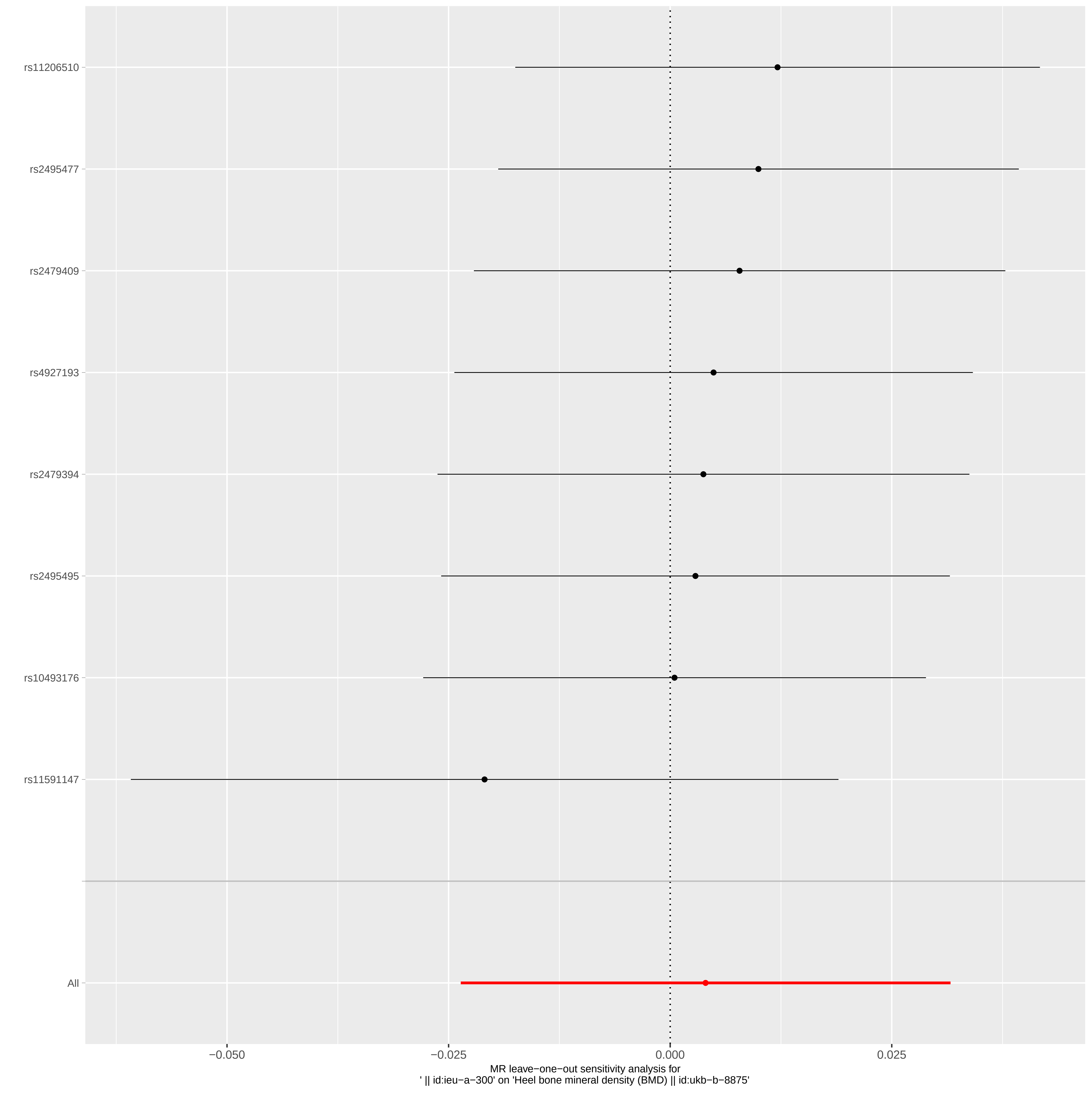
rs11206510
rs2495477
rs2479409
rs4927193
rs2479394
rs2495495
rs10493176
rs11591147
All
−0.050
−0.025
0.000
0.025
MR leave−one−out sensitivity analysis for
' || id:ieu−a−300' on 'Heel bone mineral density (BMD) || id:ukb−b−8875'

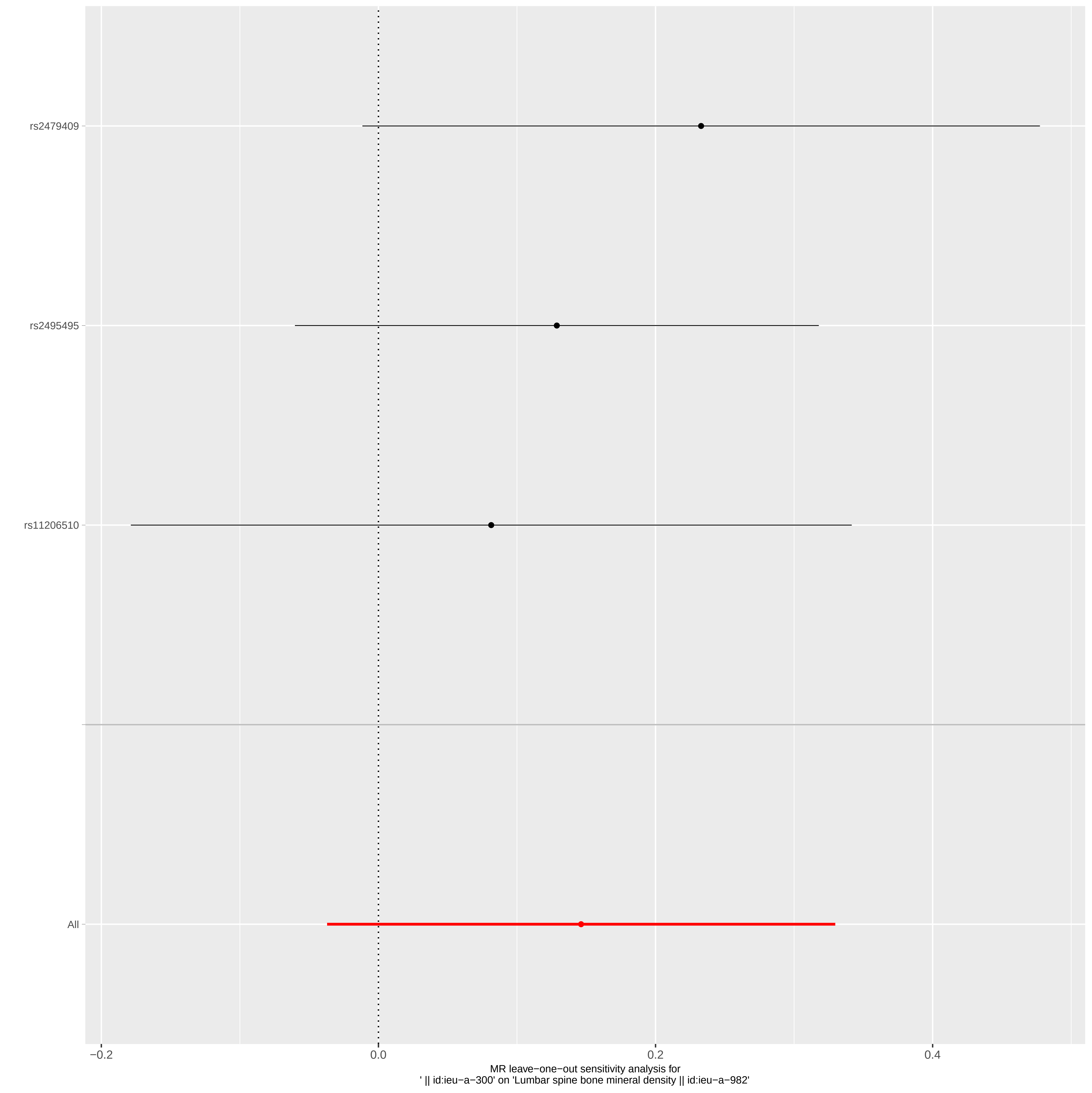
rs2479409
rs2495495
rs11206510
All
−0.2
0.0
0.2
0.4
MR leave−one−out sensitivity analysis for
' || id:ieu−a−300' on 'Lumbar spine bone mineral density || id:ieu−a−982'

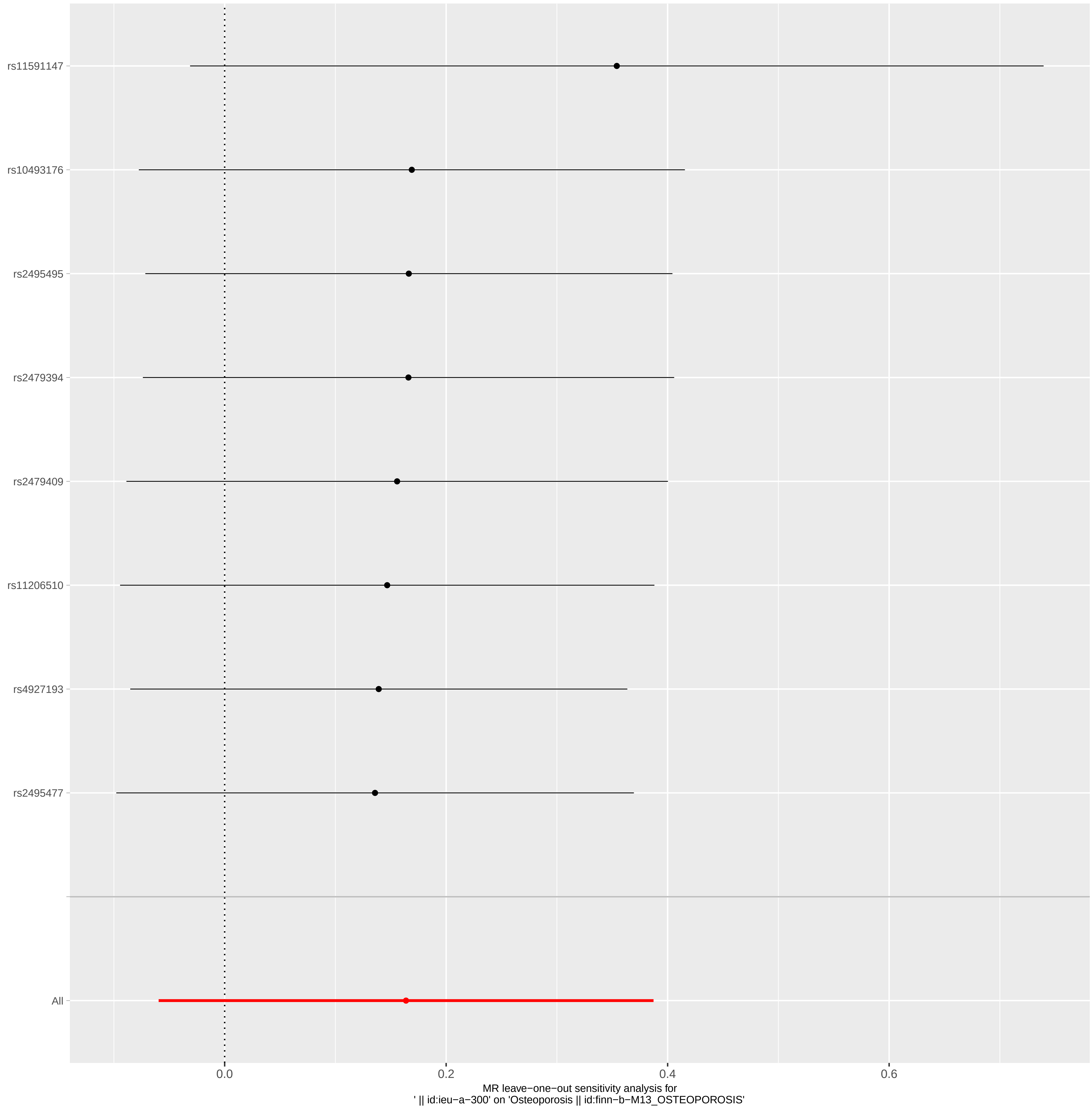

rs11591147
rs10493176
rs2495495
rs2479394
rs2479409
rs11206510
rs4927193
rs2495477
All
0.0
0.2
0.4
0.6
MR leave−one−out sensitivity analysis for
' || id:ieu−a−300' on 'Osteoporosis || id:finn−b−M13_OSTEOPOROSIS'

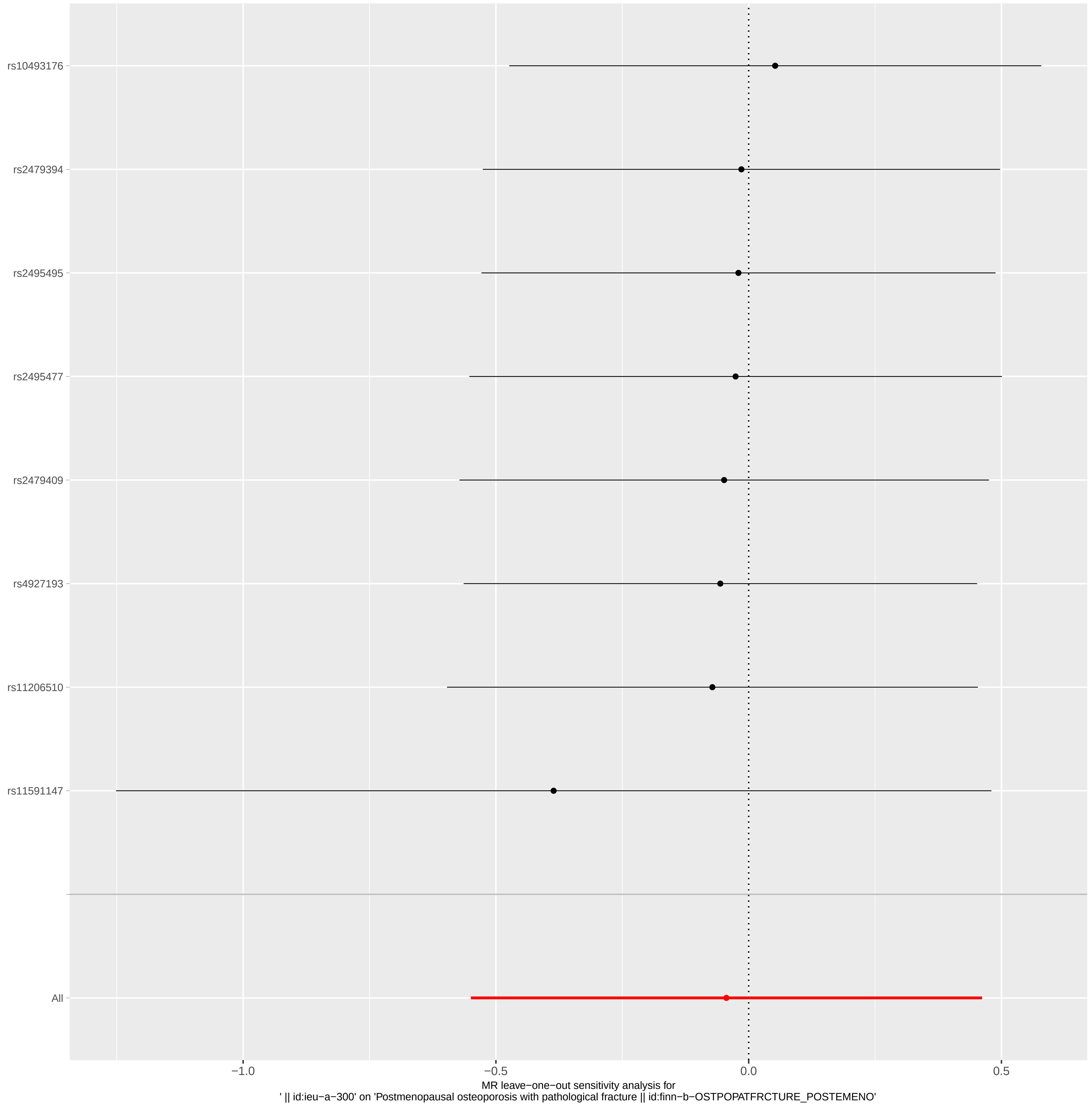

rs10493176
rs2479394
rs2495495
rs2495477
rs2479409
rs4927193
rs11206510
rs11591147
All
−1.0
−0.5
0.0
0.5
MR leave−one−out sensitivity analysis for
' || id:ieu−a−300' on 'Postmenopausal osteoporosis with pathological fracture || id:finn−b−OSTPOPATFRCTURE_POSTEMENO'

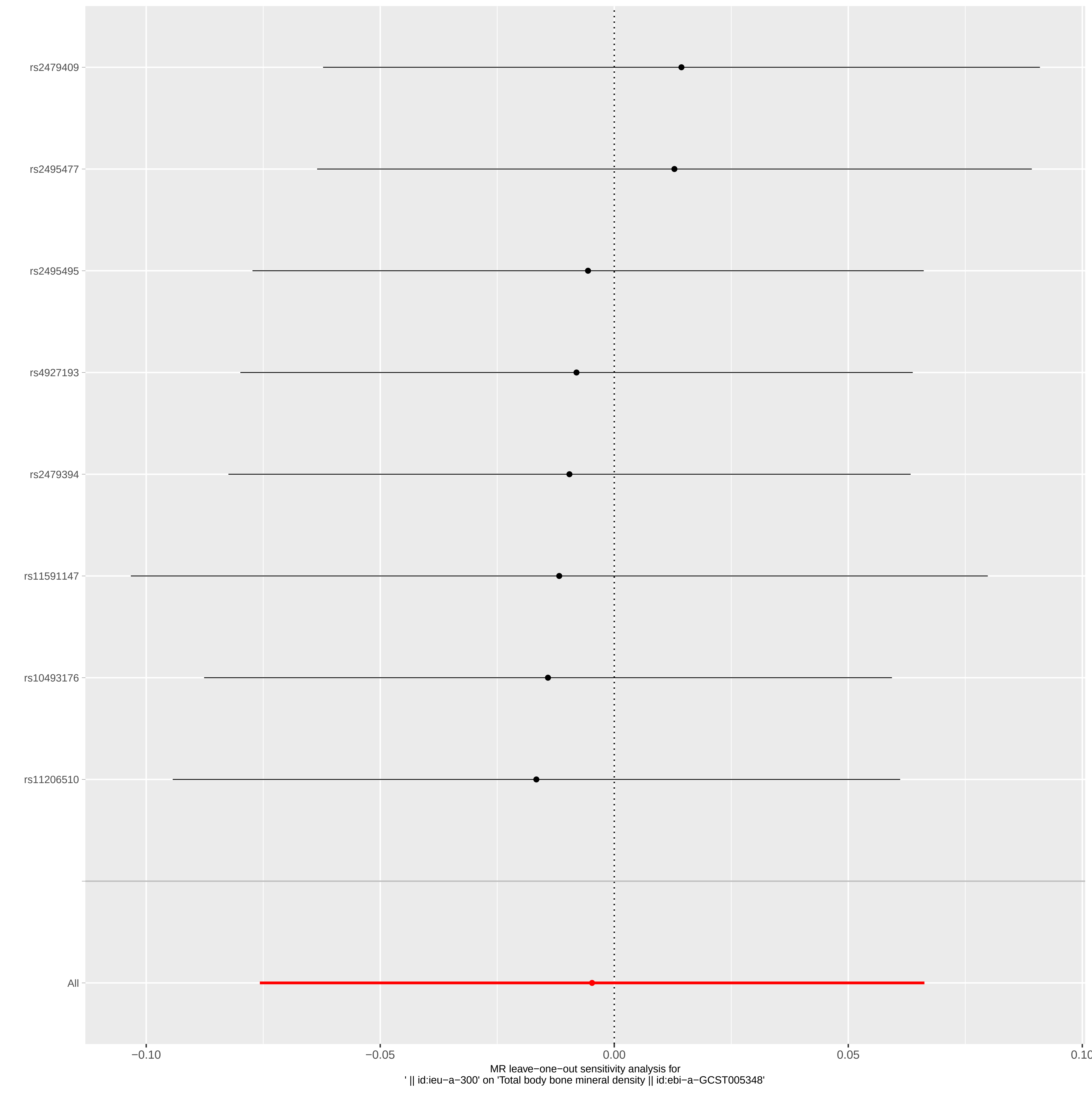

rs2479409
rs2495477
rs2495495
rs4927193
rs2479394
rs11591147
rs10493176
rs11206510
All
−0.10
−0.05
0.00
0.05
0.10
MR leave−one−out sensitivity analysis for
' || id:ieu−a−300' on 'Total body bone mineral density || id:ebi−a−GCST005348'

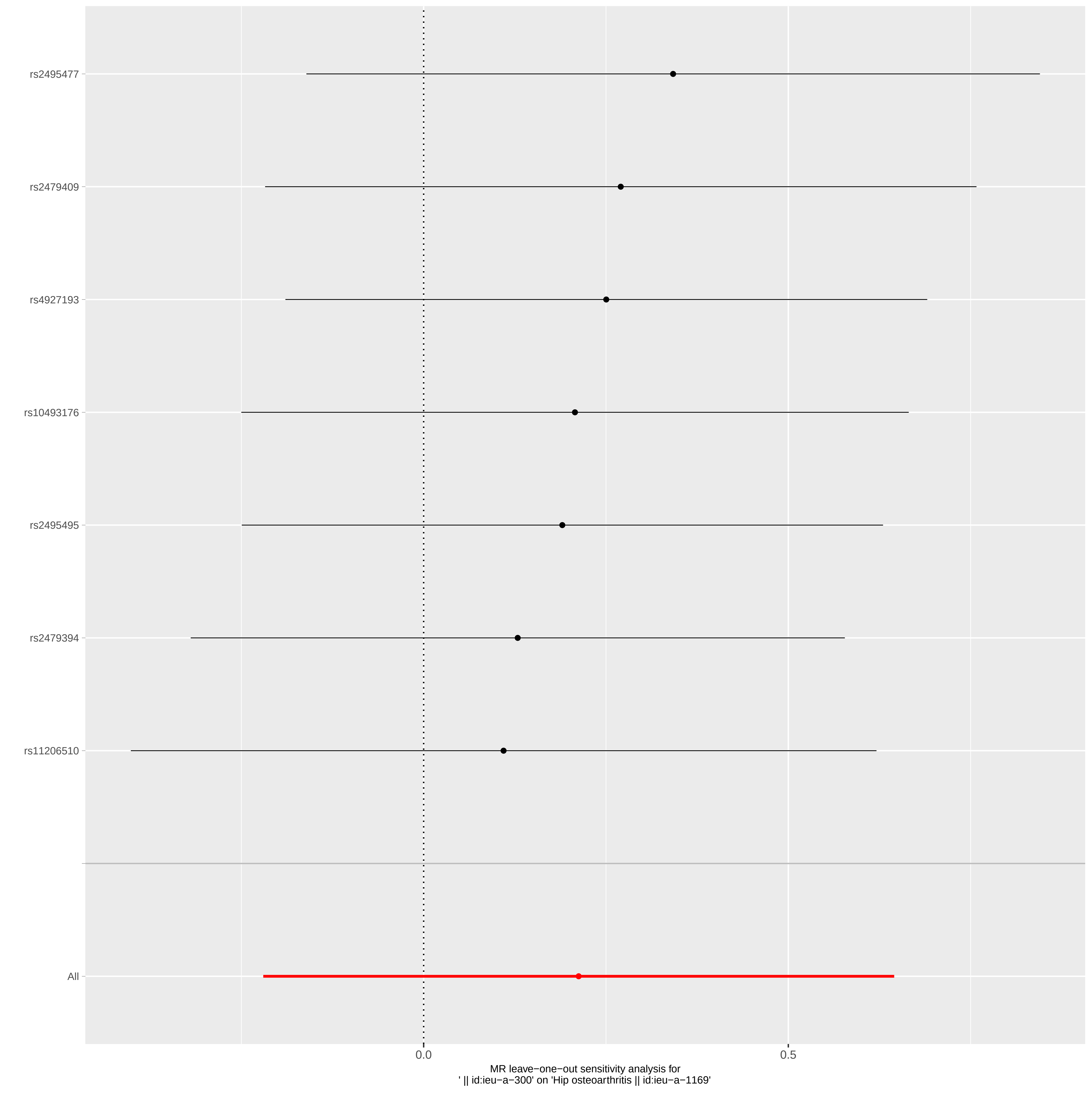

rs2495477
rs2479409
rs4927193
rs10493176
rs2495495
rs2479394
rs11206510
All
0.0
0.5
MR leave−one−out sensitivity analysis for
' || id:ieu−a−300' on 'Hip osteoarthritis || id:ieu−a−1169'

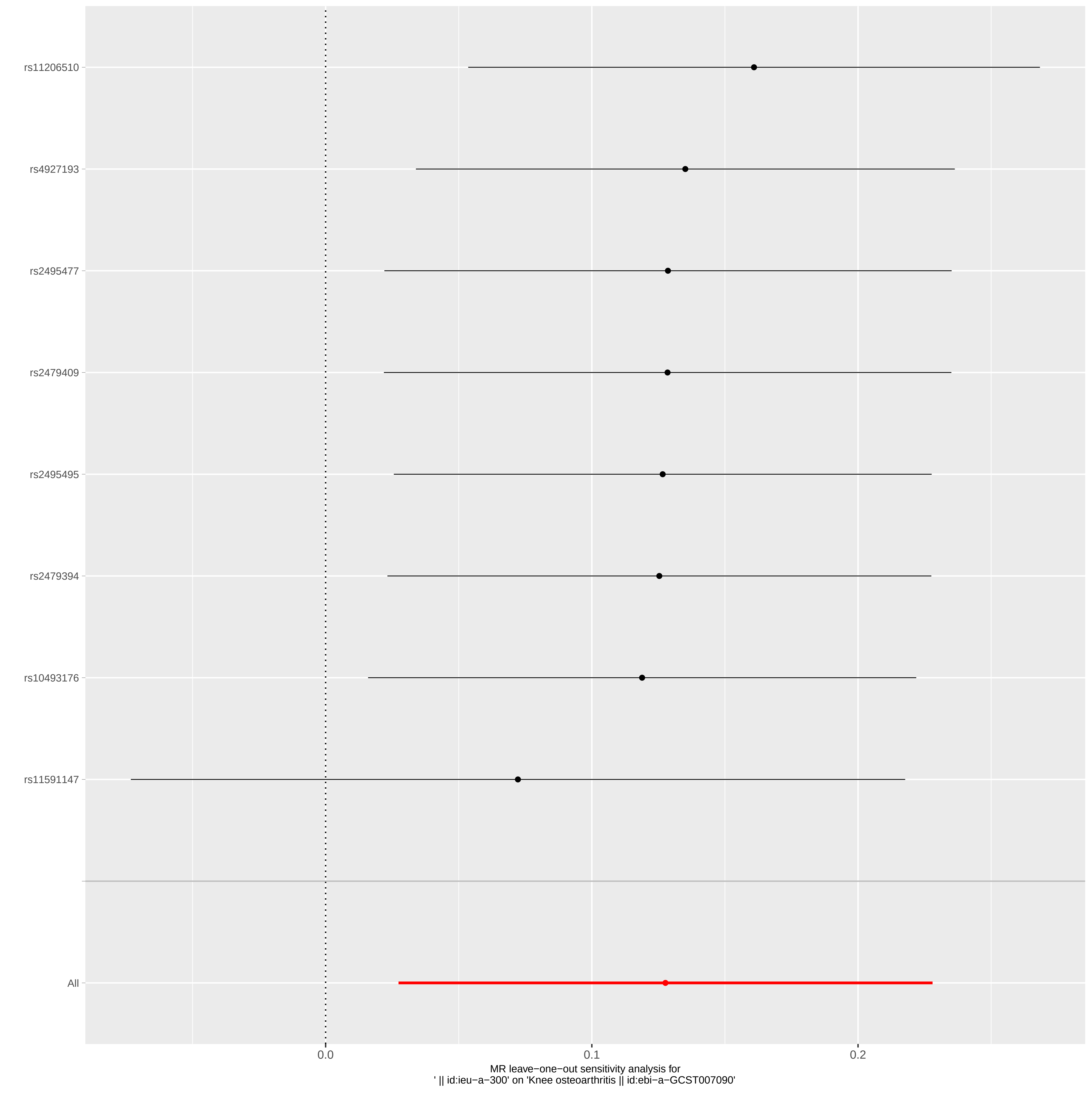

rs11206510
rs4927193
rs2495477
rs2479409
rs2495495
rs2479394
rs10493176
rs11591147
All
0.0
0.1
0.2
MR leave−one−out sensitivity analysis for
' || id:ieu−a−300' on 'Knee osteoarthritis || id:ebi−a−GCST007090'

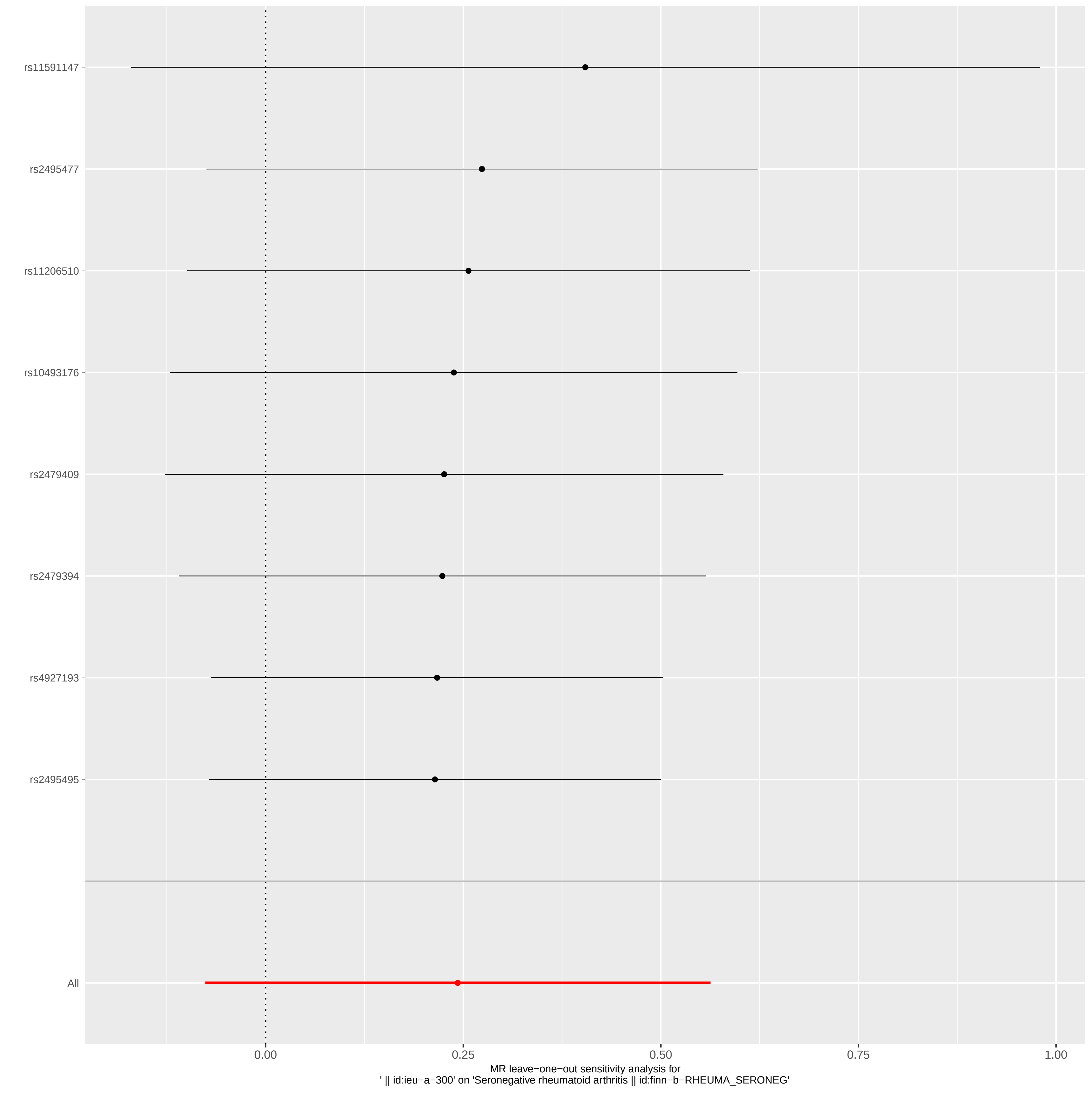

rs11591147
rs2495477
rs11206510
rs10493176
rs2479409
rs2479394
rs4927193
rs2495495
All
0.00
0.25
0.50
0.75
1.00
MR leave−one−out sensitivity analysis for
' || id:ieu−a−300' on 'Seronegative rheumatoid arthritis || id:finn−b−RHEUMA_SERONEG'

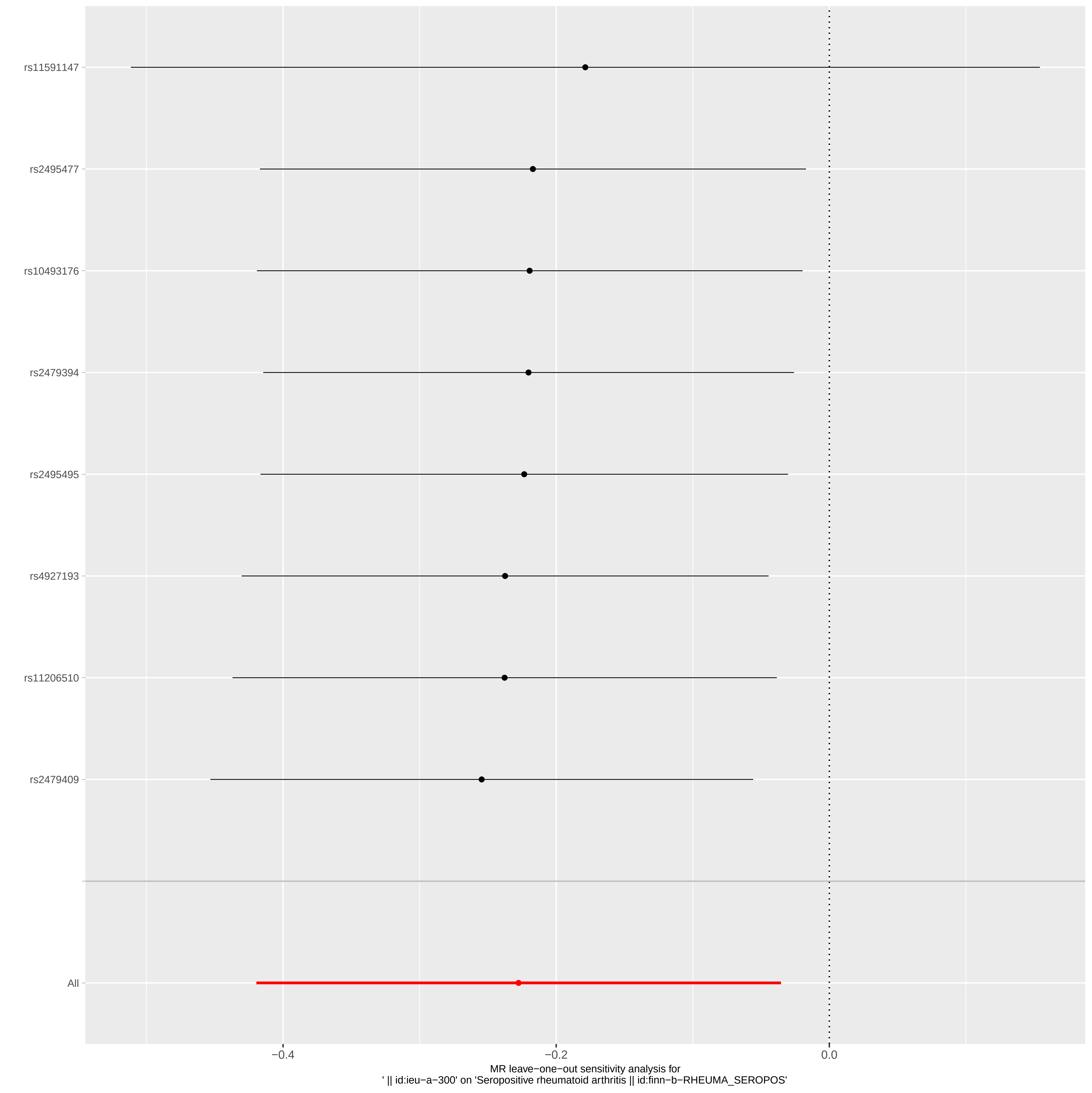
rs11591147
rs2495477
rs10493176
rs2479394
rs2495495
rs4927193
rs11206510
rs2479409
All
−0.4
−0.2
0.0
MR leave−one−out sensitivity analysis for
' || id:ieu−a−300' on 'Seropositive rheumatoid arthritis || id:finn−b−RHEUMA_SEROPOS'
